# The impact of COVID-19 vaccination on long COVID prevention in the Omicron era: An updated systematic review and meta-analysis

**DOI:** 10.64898/2026.10.12.26365288

**Authors:** Zoe Marjenberg, Michael J. Peluso, Elke Wynberg, Sultan Abduljawad

## Abstract

**Objectives:** To evaluate the impact of COVID-19 vaccination on the prevention of long COVID (LC) in the Omicron era through an updated systematic review and meta-analysis.

**Methods:** We performed an update of our previously published systematic review and meta-analysis of vaccination and LC risk (searches between 1 January 2022 and 1 March 2024). Databases (Embase, MEDLINE, PubMed, Europe PMC, LILACS, Cochrane COVID-19 Study Register) were searched for studies published between 1 March 2024 and 13 July 2026.

Observational studies assessing the impact of vaccination on the risk of LC four or more weeks after SARS-CoV-2 infection were included. Vaccines were restricted to those centrally authorised in the European Union. Random-effects meta-analyses, restricted to adults, were performed to calculate pooled odds ratios (OR) of LC in three comparisons: vaccinated (any dose) versus unvaccinated; booster vaccination versus unvaccinated; and booster vaccination versus primary course vaccination only.

**Results:** Nineteen new studies were identified, of which ten were included in the updated meta-analyses alongside eight studies from the original review. The pooled OR of LC in vaccinated adults (any dose) versus unvaccinated was 0.75 (95% confidence interval [CI] 0.67–0.84; 13 studies). ORs were also lower for booster vaccination versus unvaccinated (0.73; 95% CI 0.65–0.82; 6 studies), and booster vaccination versus primary course vaccination (0.63; 95% CI 0.53–0.75; 6 studies). Significant associations between vaccination and lower LC odds were observed in subgroup analyses of mRNA-only vaccination and LC outcomes three or more months after infection.

**Conclusions:** This updated review and meta-analysis reinforces our previous finding that vaccination may reduce the burden of LC in the Omicron era, with booster doses potentially providing additional protection. Although evidence in high-risk groups now prioritised for vaccination in most countries remains limited, these findings clearly demonstrate the benefits of vaccination and support current seasonal vaccination strategies.

## Introduction

Long COVID (LC) is an umbrella term for related, but not identical, definitions of persistent symptoms following Severe Acute Respiratory Syndrome Coronavirus 2 (SARS-CoV-2) infection.^1–5^ This debilitating condition reduces individuals’ quality of life and places a considerable burden on public health services.^6–8^

We previously reported a systematic review and meta-analysis assessing the impact of COVID-19 vaccination, including booster vaccination, on the risk of LC following SARS-CoV-2 infection during the Omicron era (PROSPERO CRD42024501445).^9^ In total, 31 observational studies published between 1 January 2022 and 1 March 2024 were identified, 11 of which were suitable for pairwise meta-analyses. The pooled odds ratio (OR) of LC (symptoms ≥4 weeks after infection) in vaccinated individuals (any dose number per study definitions, restricted to EU-authorised vaccines at the time) versus unvaccinated was 0.77 (95% confidence interval [CI] 0.70–0.85; 10 studies). ORs were lower for booster versus unvaccinated (OR 0.74; 95% CI 0.63–0.86; 4 studies), and booster versus primary course vaccination (OR 0.77; 95% CI 0.65–0.92; 3 studies).^9^

This update was undertaken to provide a more current estimate of the impact of vaccination on Omicron-era LC risk, and to assess whether a larger evidence base would support subgroup analyses. We present these updated results here, extending our original search to studies published or released as pre-prints between 1 March 2024 and 13 July 2026.

## Methods

This update followed the same search strategy and study inclusion methodology as the original review,^9^ except that: (i) only studies assessing the association between vaccination and LC risk were eligible (studies reporting prevalence of LC or vaccination association with LC severity were not captured); and (ii) child/adolescent-only studies were excluded from the meta-analyses. Dates of the update searches were 1 March 2024 to 13 July 2026.

Three previously performed meta-analyses were updated: vaccinated (any dose) versus unvaccinated, booster versus unvaccinated, and booster versus primary course vaccination. The meta-analysis feasibility assessment is detailed in the Supplementary materials. For each meta-analysis, subgroup analyses were performed restricted to (i) studies of mRNA vaccines only and (ii) studies ascertaining LC outcomes three or more months after SARS-CoV-2 infection (described hereafter as post-COVID-19 condition [PCC]) to more closely align with current definitions which require symptoms to persist for approximately three months.^1,2,4^ Sensitivity analyses, including an additional post-hoc sensitivity analysis comparing booster vaccination to no booster vaccination (0–2 doses), are described in the Supplementary materials.

## Results

The updated search identified 21 eligible publications (19 peer-reviewed articles and two pre-prints), representing 19 new studies and one secondary analysis of a study identified in the original review^10^ (Supplementary Figure 1; Supplementary Table 1). Risk of bias (Newcastle-Ottawa Scale or Joanna Briggs Institute) was low in 14 studies and medium in five (Supplementary Table 2). Study design and SARS-CoV-2 infection characteristics are summarised in Supplementary Tables 2 and 3.

Vaccination comparisons in the update studies included: vaccinated versus unvaccinated (dose definitions varied; 12 studies); booster (≥3 doses) versus unvaccinated (four studies); booster versus primary course (four studies); and recent (within the six months prior to infection, or a stated seasonal period) versus no recent vaccination (two studies). The secondary analysis compared booster with no booster (0–2 doses).^10^ Vaccination characteristics are presented in Supplementary Table 4.

Definitions of LC employed by each study are summarised in Supplementary Table 5. Minimum assessment times after SARS-CoV-2 infection in the update studies included four weeks (seven studies), six weeks (one study), two months (three studies), three months (seven studies), and six months (one study). Sixteen studies reported associations with composite LC risk and seven reported with individual symptoms. Controlled variables and composite LC outcome associations are presented in Supplementary Tables 6 and 7, respectively.

Data included in meta-analyses are shown in Supplementary Table 8. The updated vaccinated-versus-unvaccinated meta-analysis included 13 studies (six new and seven retained from the original analysis after excluding three child/adolescent-only studies). Vaccination was associated with significantly lowered pooled LC odds than unvaccinated individuals (OR 0.75 [95% CI 0.67–0.84]; Figure 1A), consistent with our previous adult sensitivity analysis (OR 0.79 [95% CI 0.72–0.87]).^9^ Between-study heterogeneity was considerable (*I*^2^=87.3%). Subgroup ORs were 0.63 (95% CI 0.51–0.78; Figure 1B) for mRNA-only vaccination and 0.77 (95% CI 0.61–0.98; Figure 1C) for PCC. No sensitivity analysis lost significance from the main analysis (Supplementary Table 9). The Egger’s test was non-significant and the funnel plot was generally symmetrical (Supplementary Figure 2), indicating no evidence of publication bias.

**Figure 1.**
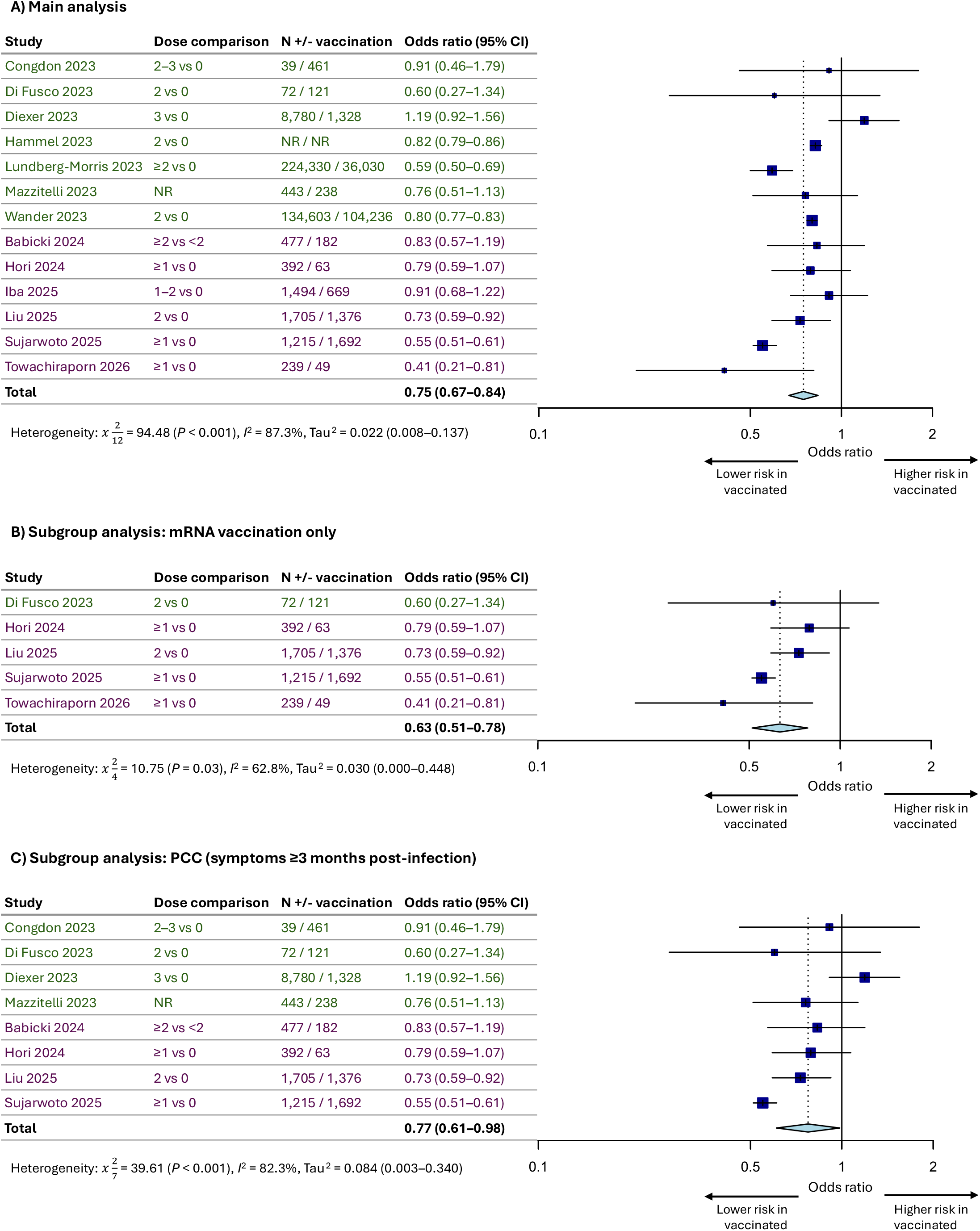
Forest plot for the effect of COVID-19 vaccination on the odds of LC compared with unvaccinated individuals. Original search studies = green; updated search studies = purple. CI, confidence interval; PCC, post-COVID-19 condition.

**Figure 2.**
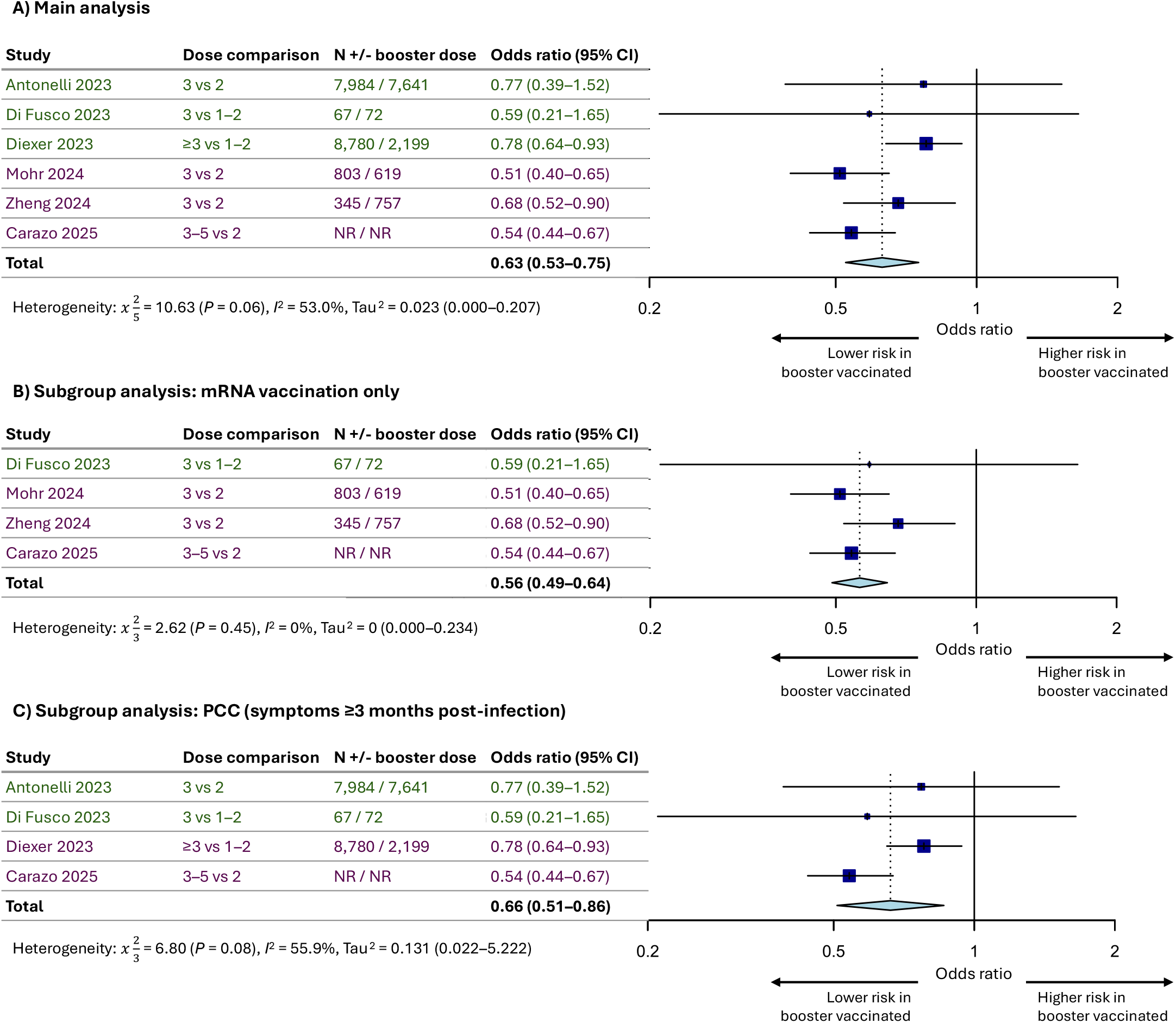
Forest plot for the effect of COVID-19 booster doses on the odds of LC, compared with primary course vaccinated individuals. Original search studies = green; updated search studies = purple. CI, confidence interval; PCC, post-COVID-19 condition.

Booster vaccination was associated with significantly lowered pooled LC odds than primary course vaccination (OR 0.63 [95% CI 0.53–0.75]; three new studies and three retained from the original analysis; Figure 2A), lower than our previous analysis (0.77 [95% CI 0.65– 0.92]).^9^ Between-study heterogeneity was substantial (*I*^2^=53.0%). The association between booster vaccination and reduced odds remained significant in the mRNA-only (OR 0.56 [95% CI 0.49–0.64]; Figure 2B) and PCC (four studies OR 0.66 [95% CI 0.51–0.86; Figure 2C) subgroup analyses. Updated booster-versus-unvaccinated results (OR 0.73 [95% CI 0.65–0.82]; two new studies and four retained from the original analysis) are presented in Supplementary Figure 3. No sensitivity analysis for either comparison lost significance from the main analysis (Supplementary Tables 10 and 11).

A sensitivity analysis pooling booster-versus-no-booster outcomes from 11 studies (Supplementary Figure 4) gave similar ORs (0.66 [95% CI 0.62–0.71]), including for the mRNA (0.57 [95% CI 0.50–0.65]) and PCC (0.66 [95% CI 0.57–0.77]) subgroup analyses.

Certainty of evidence for all meta-analyses, assessed using Grading of Recommendations Assessment, Development, and Evaluation (GRADE), was very low for all meta-analyses due to observational study design, serious inconsistency, and serious indirectness (Supplementary Table 12).

## Discussion

This update reinforces the principal finding of our original analysis: COVID-19 vaccination before SARS-CoV-2 infection in the Omicron era is associated with a significantly reduced risk of LC. Moreover, new subgroup analyses demonstrated that mRNA vaccines were more strongly associated with lower LC risk compared with the corresponding main analyses, with markedly lower heterogeneity. As mRNA vaccines account for most doses administered,^8,11^ these estimates may better reflect the protection expected under current vaccination strategies.

Definitions of LC have evolved, with PCC now used to define symptoms persisting for at least three months post-infection.^1,2,4,5^ Restriction to studies assessing PCC reduced the potential inclusion of transient post-acute symptoms and improved temporal alignment with current definitions. Comparable estimates in these subgroups suggest that the reduced risk of LC associated with vaccination was not driven by studies using earlier assessment thresholds. Alignment with current definitions nonetheless remains partial, as few studies specified a required symptom duration, applied a functional-impairment threshold, or excluded alternative diagnoses, as consensus definitions require.

Interpretation of these findings should also consider changes in COVID-19 vaccination policy. Since approximately 2024, most countries have offered seasonal vaccinations, mainly to high-risk groups, and uptake has been modest.^12,13^ Evidence specific to populations currently prioritised for seasonal vaccination remains limited, and further studies including contemporary variants, high-risk populations, and assessing impact of seasonal vaccination are needed.

The limitations of this evidence base (observational designs, variable LC definitions and vaccination status classifications, residual confounding, and collider bias) were detailed in the original publication^9^ and apply equally here, and certainty of evidence was very low across all analyses. Because seasonal vaccination now predominantly targets high-risk groups, confounding by underlying risk and health-seeking behaviour may be greater in this population than in the included studies.

In conclusion, COVID-19 vaccination was associated with a reduced risk of LC in the Omicron era, with further risk reductions associated with additional doses. The impact of vaccination remained consistent in subgroup analyses restricted to studies assessing PCC and was stronger in analyses restricted to mRNA vaccines, supporting the robustness and contemporary relevance of the overall findings. These data provide further supportive evidence for seasonal COVID-19 vaccination policies and may inform the development of evidence-based guidance on LC prevention.

## Data Availability

All data produced in the present work are contained in the manuscript or supplementary

## Acknowledgements

The authors would like to thank Dr Amanda Gallagher (BioNTech SE) for editorial support, Ms Kate Misso (Maverex Ltd) for designing and running the search strategies, and Dr Daniel Mitchell (Maverex Ltd) for statistical support.

