## Supplementary for "The impact of COVID-19 vaccination on long COVID prevention in the Omicron era: An updated systematic review and meta-analysis"

### Supplementary tables

Supplementary Table 1. Publications identified by the original and updated SLR

**Publication/pre-print**

**Original SLR publications (up to March 2024)***

AlBahrani 2023^1^

Antonelli 2023^2^

Brown 2023^3^

Congdon 2023^4^

Di Fusco 2023^5^

Diexer 2023^6^

Gallant 2023^7^

Hammel 2023^8^

Herting 2023^9^

Li 2023^10^

Lundberg-Morris 2023^11^

Mazzitelli 2023^12^

Mikolajczyk 2023^13^

Spiliopoulos 2023^14^

Richard 2023^15^

Wander 2023^16^

Woldegiorgis 2023^17^

Huh 2024^18^

Razzaghi 2024^19^

Wu 2024^20^

**New SLR publications (March 2024 to July 2026)**

Babicki 2024^21^

Hori 2024^22^

Iba 2024^23^ (same study as Iba 2025)

Malden 2024^24^

Mohr 2024^25^

Omori 2024^26^

Sun 2024^27^ (secondary analysis of Di Fusco 2023)

Xie 2024^28^

Zheng 2024^29^

Butzin-Dozier 2025^30^ (pre-print)

Carazo 2025^31^

Caspersen 2025^32^

Iba 2025^33^ (same study as Iba 2024)

Liu 2025^34^

Lutz 2025^35^

Rai 2025^36^

Sujarwoto 2025^37^

Yadaw 2025^38^ (pre-print)

Yousaf 2025^39^

Lin 2026^40^

Towachiraporn 2026^41^

* Only studies reporting vaccination association with risk of long COVIDSLR, systematic literature review.

Supplementary Table 2. Study characteristics of all included SLR studies from both the initial and updated searches

| **Study** | **Country** | **Data source** | **Population (age limits, years)** | **Population age** | **Study design** | **N Omicron participants** | **Risk of bias** |
| --- | --- | --- | --- | --- | --- | --- | --- |
| **Original SLR studies (up to March 2024)** | | | | | | | |
| AlBahrani 2023 | Saudi Arabia | Two medical centers | Healthcare workers (adults) | Mean 36.1 (SD 7.6) | Cross-sectional | 243 | High (JBI) |
| Antonelli 2023 | UK | COVID Symptom Study | Adults (18+) | Primary course vaccination:  Mean 44.3 (SD 17.7)  Booster vaccination:  Mean 45.5 (SD 16.3) | Case-control | 15,625 | Low (NOS) |
| Brown 2023 | Australia | Queensland Department of Health’s Notifiable Conditions System | Adults (18+) | 18–39: 26.0%  40–49: 17.5%  ≥50: 56.6% | Prospective | 2,195 | Medium (NOS) |
| Congdon 2023 | USA | Montefiore Medical Center and Montefiore Medical Group | Adult patients meeting criteria to be prescribed nirmatrelvir/ritonavir (18+) | Mean 50.6 (SD 15.2) | Retrospective | 500 | Low (NOS) |
| Di Fusco 2023 | USA | Consumer Value Stores (CVS) Health United States test sites | Adult outpatients (18+) | Mean 42.0 (SD 14.5) | Prospective | 328 | Medium (NOS) |
| Diexer 2023 | Germany | DigiHero (population-based cohort study on digital health research) | Adults (18+) | *18-29: 13.5%*  *30-39: 21.0%*  *40-49: 20.1%*  *50-59: 22.4%*  *60-69: 14.1%*  *≥70: 4.8%* | Prospective | 11,560 | Low (NOS) |
| Gallant 2023 | Canada | Biobanque Québécoise de la COVID-19 (BQC19) | Adults | Mean 41.5 (range 19.2–87.7) | Prospective | 290 | Low (NOS) |
| Hammel 2023 | USA | US Veterans Affairs health care system | Military veterans (adults) | 56.7 (16.3); Median 58 (IQR 43–71) | Retrospective | 245,857 (including Delta) | Medium (NOS) |
| Herting 2023 | Germany | Liver transplant center | Adult liver transplant patients (18+) | Median 56 (IQR 42–65) | Prospective | 80 | Medium (NOS) |
| Li 2023 | Singapore | Online survey | Children and young people (0–18) | Median 6.0 (IQR 3.0–10.0) | Cross-sectional | 471 | Low (JBI) |
| Lundberg-Morris 2023 | Sweden | Swedish healthcare registries | Adults (18+) | *Vaccinated:*  *Median 42 (IQR 32–53)*  *Unvaccinated:*  *Median 39 (IQR 29–50*) | Retrospective | 260,390 | Low (NOS) |
| Mazzitelli 2023 | Italy | Infectious and Tropical Disease Unit of Padua University Hospital, Padua | Adult outpatients | Median 66 (IQR 54–77) | Retrospective | 681 | Low (NOS) |
| Mikolajczyk 2023 | Germany | German National Cohort (NAKO Gesundheltsstudie) | Adults (20+) | *20-29: 3.8%*  *30-39: 14.5%*  *40-49: 19.4%*  *50-59: 32.8%*  *60-69: 9.1%*  *≥70: 9.1%* | Cross-sectional | 45,760 | Low (JBI) |
| Spiliopoulos 2023 | Denmark | Danish Nationwide Questionnaire Study (EFTER-COVID) | Adults (15+) | Median 54 (IQR 42–64) | Prospective | 13,724 | Low (NOS) |
| Richard 2023 | USA | Voluntary recruits (active-duty military) | Active-duty US military health system beneficiaries (adults) | *18-29: 31.1%*  *30-39: 43.1%*  *≥40: 25.8%* | Prospective | 853 | Low (NOS) |
| Wander 2023 | USA | US Veterans Affairs | Military veterans (adults) | Mean 61.4 (SD 16.1) | Retrospective | 388,980 | Medium (NOS) |
| Woldegiorgis 2023 | Australia | Western Australia Department of Health | Adult population-based cohort (18+) | 18-29: 20.9%  30-39: 21.0%  40-49: 18.7%  50-59: 18.0%  60-69: 11.6%  ≥70: 9.7% | Prospective | 11,697 | Low (NOS) |
| Huh 2024 | South Korea | A combined COVID-19 database constructed by the Korea Disease Control and Prevention Agency and the National Health Insurance Service of the Republic of Korea | Adults (18+) | Vaccinated:  Mean 43.9 (SD 16.31)  Unvaccinated:  Mean 43.7 (SD 17.43) | Retrospective | 7,998,854 | Low (NOS) |
| Razzaghi 2024 | USA | Electronic health record data from 41 US health systems | Children (5 to <18) | *Vaccinated:*  *Mean 12.4 (SD 3.58)*  *Unvaccinated:*  *Mean 11.1 (SD 3.66)* | Retrospective | 289 | Low (NOS) |
| Wu 2024 | USA | National Institute of Health Researching COVID to Enhance Recovery (RECOVER) Initiative | Children (5–11) and adolescents (12–20) | Children:  Median 8 (IQR 6–10)  Adolescents:  Median 15 (IQR 13–17) | Retrospective | 273,629 | Medium (NOS) |
| **New SLR studies (March 2024 to July 2026)** | | | | | | | |
| Babicki 2024 | Poland | STOP-COVID registry | Adults (18+) | Mean 55.7 (SD14.0) | Retrospective | 659 | Medium (NOS) |
| Hori 2024 | Japan | Self-reported questionnaire of individuals in Yao City | Adults (18–79) | Mean 44.4 (SD 14.6) | Cross-sectional | 3,552 | Low (JBI) |
| Iba 2024 & Iba 2025 | Japan | Self-reported questionnaire of individuals in Shinagawa City | Adults (20–69) | Mean 42.3 (SD 11.6) | Cross-sectional | Iba 2024: 8,392  Iba 2025: 7,936 | Low (JBI) |
| Malden 2024 | USA | Vaccine Safety Datalink network EHRs | Any | *Mean 37.1 (SD 17.9)* | Retrospective | 216,246 | Low (NOS) |
| Mohr 2024 | USA | Multicentre Preventing Emerging Infections through Vaccine Effectiveness Testing Project (Project PREVENT) | Adults (18+) | Primary course vaccination:  18–29: 26.0%  30–39: 29.6%  40–49: 22.1%  50–64: 21.5%  ≥65: 0.8%  Booster vaccination:  18–29: 20.0%  30–39: 35.6%  40–49: 20.4%  50–64: 21.8%  ≥65: 2.1% | Prospective | 1,422 | Medium (NOS) |
| Omori 2024 | Japan | Self-reported questionnaire administered to patients attending the fever outpatients clinic at the department of Respiratory Medicine, Kuramochi Clinic Interpark, Ustunomiya, Tochigi | Any | Mean 40.5 (SD 9.5) | Cross-sectional | 3,326 | Low (JBI) |
| Xie 2024 | USA | Healthcare databases of the US Department of Veterans Affairs | Adults (18+) | *Vaccinated:*  *Mean 58.24 (SD 15.08)*  *Unvaccinated:*  *Mean 58.38 (SD 15.26)* | Retrospective | 125,310 | Low (NOS) |
| Zheng 2024 | Hong Kong | Self-reported online questionnaire | Adults (18+) | 18–30: 51.3%  31–40: 22.4%  41–50: 13.4%  51–60: 8.2%  >60: 4.7% | Cross-sectional | 1,542 | Low (JBI) |
| Butzin-Dozier 2025 | USA | National COVID Cohort Collaborative (N3C) | Any | Vaccinated:  0–17: 37%  17–49: 29%  49–70: 22%  70–107: 2%  Unvaccinated:  0–17: 51%  17–49: 34%  49–70: 13%  70–107: 1% | Retrospective | 519,980* | Low (NOS) |
| Carazo 2025 | Canada | Self-reported questionnaire administered to healthcare workers in Quebec, laboratory registry containing all SARS-CoV-2 NAATs | Adults, healthcare workers (18+) | LC cases:  18–39: 29.7%  40–49: 35.1%  50–59: 25.4%  ≥60: 9.8%  COVID-19 controls:  18–39: 38.6%  40–49: 32.0%  50–59: 21.1%  ≥60: 8.4% | Retrospective | 7,788* | Medium (NOS) |
| Caspersen 2025 | Norway | Self-reported questionnaire of individuals in four cohorts: The Norwegian Mother, Father, and Child Cohort Study, The Norwegian Influenza Pregnancy Cohort, The Senior Cohort, and The Young Adult Cohort | Adults and adolescents (11–80) | NR | Cross-sectional | 26,701 | Low (JBI) |
| Liu 2025 | USA | Kaiser Permanente Southern California (KPSC) integrated healthcare system electronic medical record database | Adults (18+) | 18–45: 30.5%  45–55: 22.6%  ≥55: 46.9% | Case-control | 5,040 | Low (NOS) |
| Lutz 2025 | USA | Self-reported questionnaire and EHRs of Indigenous individuals in Chinle, Arizona and Tuba City, Arizona (both in the Navajo Nation), and in Whiteriver, Arizona in the White Mountain Apache Tribal lands | Adults and children from rural Indigenous communities | Median 48.9 (IQR 36.6–59.5) | Prospective | 145 | Low (NOS) |
| Rai 2025 | UK | The Health Improvement Network (THIN) | Adults (18+) | Highest risk cohort, no seasonal COVID-19 vaccination:  50–<65: 2.0%  75–<85: 60.1%  ≥85: 35.0%  Highest risk cohort, seasonal COVID-19 vaccination:  65–<75: 1.8%  75–<85: 61.9%  ≥85: 35.1%  High risk cohort, no seasonal COVID-19 vaccination:^†^  18–<50: 25.8%  50–<65: 53.1%  65–<75: 21.1%  75–<85: 67.5%  ≥85: 25.8%  High risk cohort, seasonal COVID-19 vaccination:^†^  18–<50: 8.4%  50–<65: 54.0%  65–<75: 37.6%  75–<85: 61.5%  ≥85: 8.4%  Non-high/highest-risk cohort: NR | Retrospective | 8,507 | Low (NOS) |
| Sujarwoto 2025 | Indonesia | 33 community health centres and hospitals in Malang Regency, Indonesia | Adults (18–99) | Mean 43.48 (SD 15) | Cross-sectional | 5,735 (including non-EMA vaccine recipients) | Low (JBI) |
| Yadaw 2025 | USA | National COVID Cohort Collaborative Enclave | Any (1+) | *1-20: 16%*  *20-40: 27%*  *40-65: 37%*  *>65: 21%* | Retrospective | 549,670 | Medium (NOS) |
| Yousaf 2025 | USA | Paediatric Research Observing Trends and Exposures in COVID-19 Timelines (PROTECT) study | Children and adolescents (5–17) | LC cases:  Median 10.0 (IQR 7.0–11.9)  COVID-19 controls:  Median 10.3 (IQR 7.8–12.7) | Case-control |  | Low (NOS) |
| Lin 2026 | USA | VISION study | Adults | Vaccinated >6 months:  18–34: 25%  35–49: 38%  50–64: 25%  ≥65: 13%  Vaccinated ≤6 months:  18–34: 15%  35–49: 26%  50–64: 26%  ≥65: 32% | Prospective | 2,960 | Low (NOS) |
| Towachiraporn 2026 | Thailand | Maharaj Nakorn Chiang Mai Hospital | Adults (20+) | mRNA vaccine:  Mean 40.8 (SD 11.2)  No mRNA vaccine:  Mean 35.9 (SD 10.9) | Prospective | 288 | Medium (NOS) |

EMA, European Medicines Agency; IQR, interquartile range; JBI, Joanna Briggs Institute; LC, long COVID; NAAT, Nucleic acid amplification test; NOS, Newcastle-Ottawa Scale; SD, standard deviation; SLR, systematic literature review.
*Italics = age only reported for the entire population including non-Omicron variants.*
* Relevant outcomes only reported for these cohorts.
† Percentages do not total 100%, potential publication error.

Supplementary Table 3. Infection characteristics of all included SLR studies from both the initial and updated searches

| **Study** | **Infection period (Omicron only)** | **Omicron subvariants reported** | **Infection confirmation** | **Severity of acute disease** |
| --- | --- | --- | --- | --- |
| **Original SLR studies (up to March 2024)** | | | | |
| AlBahrani 2023 | May 2022 to Aug 2022 | NR | PCR test | NR |
| Antonelli 2023 | Dec 20 2021 to Apr 1 2022 | NR | PCR or antigen test | - Hospitalised: 2% |
| Brown 2023 | Jun 12 2022 to Jun 25 2022 | NR | PCR test | NR |
| Congdon 2023 | Dec 29 2021 to Apr 2022 | NR | PCR test | - Hospitalised: 1% |
| Di Fusco 2023 | Jan 31 2022 to Apr 30 2022 | BA.1, BA.2 | PCR or antigen test | NR |
| Diexer 2023 | Dec 21 2021 onwards | NR | PCR test, >1 antigen test, or 1 antigen test with symptoms | NR |
| Gallant 2023 | Dec 2021 to Apr 2022 | NR | PCR test | - Hospitalised: 1.4% |
| Hammel 2023 | Dec 1 2021 to Feb 2 2022 | NR | PCR or antigen test | NR |
| Herting 2023 | Dec 20 2021 to Jul 31 2022 | BA.1, BA.2, BA.4, BA.5 | PCR test or antigen test with a positive nucleocapsid  antibody test after the infection or via an antigen test with typical symptoms  with a known source of infection | - Hospitalised: 8.8% |
| Li 2023 | Jan 2022 onwards | BA.1, BA.2, BA.4, BA.5, XBB | PCR or antigen test | NR |
| Lundberg-Morris 2023 | Jan 2022 to 9 Feb 2022 | NR | PCR test or ICD-10 diagnosis code | - Hospitalised: 2.7% - Intensive care: 0.08% |
| Mazzitelli 2023 | Feb 9 2022 to May 31 2022 | NR | NR, but ascertainment stated to be by nasopharyngeal swab | - Hospitalised: 8.7% |
| Mikolajczyk 2023 | Jan 2022 onwards | NR | Self-reported | NR |
| Spiliopoulos 2023 | Dec 28 2021 to Jan 15 2022 | NR | PCR test | NR |
| Richard 2023 | Jan 1 2022 to Apr 30 2022 | NR | PCR test, self-reported positive respiratory swab test, or qPCR on research swabs | - Hospitalised: 2.7% |
| Wander 2023 | Oct 1 2021 to Jan 31 2023 | NR | PCR test or ICD-10 diagnosis code | - Hospitalised: 10.3% |
| Woldegiorgis 2023 | Jul 16 2022 to Aug 3 2022 | NR | PCR or antigen test | NR |
| Huh 2024 | Jan 16 2022 to Mar 31 2022 | BA.1, BA.2 | “Test-confirmed” | - Severe: 0.6% - Critical: 0.05% |
| Razzaghi 2024 | Nov 30 2021 onwards | NR | PCR or antigen test or ICD-10 diagnosis code | NR |
| Wu 2024 | Jan 1 2022 to Nov 30 2022 | NR | PCR, serology, or antigen tests | NR |
| **New SLR studies (March 2024 to July 2026)** | | | | |
| Babicki 2024 | Jan 2022 to Dec 2022 | NR | PCR or antigen test | - Symptoms for 0–3 days: 1.2%. - Symptoms for 4–7 days: 32.6%. - Symptoms for 8–14 days: 34.6%. - Treated at home with symptoms lasting ≥14 days, a fever greater than 38°C, dyspnoea and oxygen saturation <94% for ≥3 days: 18.4%. - Hospitalised patients: 13.2%. |
| Hori 2024 | Jan 2022 to Apr 2022 | NR | PCR, antigen test, or clinical diagnosis (for symptomatic close contacts) | - Asymptomatic: 6.6% - Mild: 87.1% - Moderate: 0.8% - Severe: 0.3% - Unknown: 0.2% |
| Iba 2024 & Iba 2025 | Jul 1 2022 to Aug 31 2022 | BA.5 | PCR, antigen test, or clinical diagnosis (for symptomatic close contacts) | Iba 2024:   - Asymptomatic: 2.7% - Mild: 96.5% - Moderate/severe: 0.8%Severe: 0.3%   Iba 2025:   - PCC, Asymptomatic: 1.4% - PCC, Mild: 97.2% - PCC, Moderate: 1.4% - PCC, Severe: 0% - No PCC, Asymptomatic: 3.0% - No PCC, Mild: 96.3% - No PCC, Moderate: 0.7% - No PCC, Severe: 0% |
| Malden 2024 | Dec 1 2021 to Feb 28 2022 | NR | PCR or antigen test | NR |
| Mohr 2024 | Dec 19 2021 to Apr 27 2022 | NR | PCR or antigen test | NR |
| Omori 2024 | Jul 1 2022 to Sep 30 2022 | NR | PCR test | - Non-hospitalised: 96.1% - Hospitalised: 2.5% - Missing data: 1.4% |
| Xie 2024 | Dec 19 2021 to Jan 31 2022 | NR | Positive test (not further specified) | NR |
| Zheng 2024 | Jan 3 2022 to Apr 29 2022 | NR | Self-reported PCR or antigen test | - Asymptomatic: 4.7% - Mild: 76.2% - Moderate: 18.2% - Severe: 0.9% - Critical: 0% - Hospitalised: 0.9% - Non-hospitalised: 99.1% - ICU admission: 0% - Non-ICU admission: 100% |
| Butzin-Dozier 2025 | Dec 25 2021 to Sep 25 2022 | NR | From EHRs (not further specified) | NR |
| Carazo 2025 | Dec 19 2021 to Feb 20 2023 | BA.1, BA.2, BA.4, BA.5, BQ.1, XBB | Antigen test | NR |
| Caspersen 2025 | Jan 2022 to Feb 2022 | BA.1.1, BA.2 | PCR or antigen test, or record of laboratory confirmed infection | 11–12 years:   - Asymptomatic: 4.7% - Mild: 43.7% - Moderate: 48.7% - Severe: 2.9%   18–30 years:   - Asymptomatic: 4.6% - Mild: 40.1% - Moderate: 52.0% - Severe: 3.3%   30–65 years:   - Asymptomatic: 3.1% - Mild: 45.7% - Moderate: 48.4% - Severe: 2.8%   65–80 years:   - Asymptomatic: 2.7% - Mild: 63.5% - Moderate: 32.3% - Severe: 1.6% |
| Liu 2025 | Jan 1 2022 to Nov 30 2022 | BA.1, BA.2, BA.4, BA. 5 | SARS-CoV-2 positive lab results, ICD-10 codes, patient-reported positive home test kits, patient request for acute COVID treatment and work notes, chart review search words for COVID/SARS-CoV-2 infection, and reported care received outside of healthcare system | Long COVID:   - Hospitalised: 95.3% - Non-hospitalised: 4.7%   No long COVID:   - Hospitalised: 1.5% - Non-hospitalised: 98.5% |
| Lutz 2025 | Dec 25 2021 to Aug 31 2022 | NR | Positive test (not further specified) | - Outpatient: 89.7% - Inpatient: 10.3% |
| Rai 2025 | Sep 1 2022 to May 31 2023 | NR | Confirmed or suspected COVID diagnosis (Read code) | NR |
| Sujarwoto 2025 | Jun 15 2022 to Jun 15, 2023 | NR | Positive test (not further specified) | NR |
| Yadaw 2025 | Dec 23 2021 to Jan 4 2024 | NR | PCR, antigen, or clinical diagnosis | NR |
| Yousaf 2025 | Dec 1 2022 to May 31 2023 | NR | PCR test | Long COVID:   - Symptomatic: 86% - Asymptomatic: 14%   No long COVID:   - Symptomatic: 90% - Asymptomatic: 10% |
| Lin 2026 | Oct 5 2022 to Mar 31 2024 | NR | Positive test (not further specified) | Vaccination ≤6 months:   - None: 8.9% - Mild: 60% - Moderate: 30% - Severe: 1.2% - Non-hospitalised: 100%   No vaccination ≤6 months:   - None: 7.5% - Mild: 50% - Moderate: 39% - Severe: 3.4% - Non-hospitalised: 100% |
| Towachiraporn 2026 | Feb 1 2023 to Dec 30 2023 | NR | PCR or antigen test | All mild (population selection criteria) |

EHR, electronic health records; NR, not reported; PCC, post-COVID condition; PCR, polymerase chain reaction; qPCR, quantitative PCR.

Supplementary Table 4. Vaccination characteristics of all included SLR studies from both the initial and updated searches

| **Study** | **Vaccination status and doses reported** | **Dose number and % receiving (Omicron only)** | **Time to Omicron SARS-CoV-2 infection from last vaccine dose** | **Vaccine type (Omicron only)** |
| --- | --- | --- | --- | --- |
| **Original SLR studies (up to March 2024)** | | | | |
| AlBahrani 2023 | Unvaccinated: 0  Vaccinated: 2–4 | 0: 1.2%  2: 2.1%  3:91.8%  4: 4.9% | NR | NR |
| Antonelli 2023 | Primary course: 2  Booster dose: 3 | 2: 48.9%  3: 51.1% | NR | NR |
| Brown 2023 | No booster dose: ≤2  Booster dose: ≥3 | ≤2: 16.8%  ≥3: 83.2% | ≥6 months since last dose: 23.1% | NR |
| Congdon 2023 | Unvaccinated: 0  Primary course: 2  Booster dose: 3 | 0: 7.8%  2: 24.6%  3: 67.6% | NR | BNT162b2, mRNA-1273, Ad26.COV2.S, ChAdOx1 |
| Di Fusco 2023 & Sun 2024 | Unvaccinated: 0  Primary course: 2  Booster dose: 3 | 0: 47.3%  2: 26.2%  3: 26.5% | Primary course:   - Mean (SD) 6.9 (3.0) months   Booster dose:   - Mean (SD) 2.3 (1.9) months | BNT162b2 |
| Diexer 2023 | Unvaccinated: 0  Primary course: 1–2  Booster dose: ≥3 | 0: 10.8%  1–2: 17.9%  ≥3: 71.3% | NR | NR |
| Gallant 2023 | Unvaccinated: ≤1  Vaccinated: ≥2 | ≤1: 3.4%  ≥2: 96.6% | NR | BNT162b2, mRNA-1273 |
| Hammel 2023 | Unvaccinated: ≤2  Primary course: 2  Booster dose: 3 | NR | NR | BNT162b2, mRNA-1273, Ad26.COV2.S |
| Herting 2023 | Primary course: 2  Booster dose: 3–5 | 2: 12.2%  3: 60.2%  4: 25.6%  5: 2% | Median (IQR) 130 (88.8–183.3) days | BNT162b2, mRNA-1273, and viral vector |
| Li 2023 | Unvaccinated: ≤1  Vaccinated: ≥2 | 0–1: 56.5%  ≥2: 43.5% | NR | BNT162b2, mRNA-1273 |
| Lundberg-Morris 2023 | Unvaccinated: 0  Vaccinated: 1–5 | 0: 13.8%  1–5: 86.2% | NR | Mixed: BNT16b2, mRNA-1273, ChAd0x1 |
| Mazzitelli 2023 | Unvaccinated: NR  Vaccinated: NR | Unvaccinated: 65.1%  Vaccinated: 34.9% | NR | NR |
| Mikolajczyk 2023 | Unvaccinated: 0  Primary course: 1–2  Booster dose: 3  Additional booster dose: 4 | NR | NR | NR |
| Spiliopoulos 2023 | Unvaccinated: ≤1  Primary course: 2  Booster dose: 3 | 0: 3.5%  1: 0.6%  2: 38.4%  3: 57.5% | NR | BNT162b2, mRNA-1273, ChAd0x1, Ad26.COV2.S |
| Richard 2023 | Unvaccinated: ≤1  Primary course: 2  Booster dose: ≥3 | NR | NR | BNT162b2, mRNA-1273, Ad26.COV2.S |
| Wander 2023 | Unvaccinated: 0  Primary course: 2  Booster dose: ≥3 | 0: 26.8%  2: 34.6%  ≥3: 38.6% | NR | BNT162b2, mRNA-1273, Ad26.COV2.S |
| Woldegiorgis 2023 | No booster dose: ≤2  Booster dose: 3  Additional booster dose: ≥4 | ≤2: 7.7%  3: 75.8%  ≥4: 16.6% | NR | NR |
| Huh 2024 | Unvaccinated: 0  Primary course: 2  Booster dose: 3 | 0: 4.9%  2: 32.5%  3: 62.5% | NR | BNT162b2, mRNA-1273, ChAd0x1 |
| Razzaghi 2024 | Unvaccinated: 0  Vaccinated: ≥1 | NR | NR | mRNA |
| Wu 2024 | Unvaccinated: 0  Vaccinated: ≥1 | NR | NR | BNT162b2 |
| **New SLR studies (March 2024 to July 2026)** | | | | |
| Babicki 2024 | Unvaccinated: 0–1  Vaccinated: ≥2 | NR | NR | NR |
| Hori 2024 | Unvaccinated: 0  Vaccinated: 1–5 | 0: 11.2%  1: 0.5%  2: 27.2%  3: 36.1%  4: 22.2%  5: 2.9% | NR | BNT162b2, mRNA-1273 |
| Iba 2024 & Iba 2025 | Unvaccinated: 0  Vaccinated: ≥1 | 0: 8.2%  1: 0.6%  2: 20.0%  ≥3: 71.3% | PCC:   - <90 days: 8.5% - 90-179 days: 51.7% - ≥180 days: 29.1%   No PCC:   - <90 days: 8.6% - 90-179 days: 58.1% - ≥180 days: 25.2% | NR |
| Malden 2024 | Unvaccinated: 0  Vaccinated: ≥1 | NR | Including pre-Omicron variants:   - <90 days: 31.2% - 90–180 days: 29.7% - >180 days: 39.1% | NR |
| Mohr 2024 | Vaccinated: 3  Booster dose: 2 | NR | Booster vaccination only:   - Mean (SD): 86 (38) days - Median (IQR): 85 (61–106) days | BNT162b2, mRNA-1273 |
| Omori 2024 | Unvaccinated: 0  Partial/fully vaccinated: 1–2  Booster dose: 3  Additional booster dose: 4 | 0: 13.6%  1-2: 18.2%  3: 45.0%  4: 18.5% | NR | NR |
| Xie 2024 | Unvaccinated: 0  Vaccinated: ≥1 | NR | NR | NR |
| Zheng 2024 | Unvaccinated: 0–1  Vaccinated: 2  Booster dose: 3 | 0: 5.3%  1: 3.5%  2: 49.1%  3: 22.4% | NR | BNT162b2 |
| Butzin-Dozier 2025 | Unvaccinated: 0  Vaccinated: ≥2 | NR | NR | NR |
| Carazo 2025 | Waned immunity: 2 ≥6 months before infection  Booster dose: 3–5 | LC:   - 2: 17.5% - 3-5: 82.5%   No LC:   - 2: 11.9% - 3-5: 88.1% | 2 doses:   - Median (IQR): 8.2 (6.4–10.5) months   3–5 doses:   - Median (IQR): 7.4 (5.7–9.7) months | BNT162b2, mRNA-1273 |
| Caspersen 2025 | Unvaccinated: 0  Vaccinated: 2  Booster dose: 3 | NR | 2 doses:   - Median (IQR) 154 (141–173) days   3 doses:   - Median (IQR) 40 (26–56) | NR |
| Liu 2025 | Unvaccinated: 0  Vaccinated: 1–4 | 0: 27.3%  1: 31.4%  2: 33.8%  3: 7.0%  4: 0.4% | NR | BNT162b2, mRNA-1273 |
| Lutz 2025 | Unvaccinated: 0–1  Vaccinated: 2  Booster dose: ≥3 | 0–1: 9.7%  2: 51.7%  ≥3: 38.6% | NR | NR |
| Rai 2025 | No seasonal vaccination: 0 between Sep 1 2022 and May 31 2023  Seasonal vaccination: ≥1 between Sep 1 2022 and May 31 2023 | High risk patients:   - No seasonal vaccine: 62% - Seasonal vaccine: 38%   Highest risk patients:   - No seasonal vaccine: 40% - Seasonal vaccine: 60% | High risk patients:   - >6 months: 65.0% - <3 months: 16.8% - 3-6 months: 18.2%   Highest risk patients:   - >6 months: 37.5% - <3 months: 21.4% - 3-6 months: 41.1% | NR |
| Sujarwoto 2025 | Unvaccinated: 0  Vaccinated: ≥1 | 0: 32.0%  1: 59.5%  2: 5.8%  ≥3: 2.7% | NR | BNT162b2, mRNA-1273,  Ad26.COV2.S, ChAdOx1 |
| Yadaw 2025 | Unvaccinated: <2  Vaccinated: ≥2 | NR | NR | NR |
| Yousaf 2025 | Unvaccinated: 0  Vaccinated: 2  Booster dose: ≥3 | PCC:   - 0: 43% - 2: 43% - >2: 14%   No PCC   - 0: 23% - 2: 60% - >2: 17% | PCC:   - Median (IQR): 18 (8–30) weeks   No PCC   - Median (IQR): 20 (8–30) weeks | BNT162b2 |
| Lin 2026 | No recent vaccination: 0 ≤6 months before infection  Recent vaccination: ≥1 ≤6 months before infection | NR | ≤6 months | NR |
| Towachiraporn 2026 | No mRNA vaccination: 0  mRNA vaccinated: ≥1 | NR | NR | BNT162b2, mRNA-1273 |

IQR, interquartile range; NR, not reported; PCC, Post-COVID condition; SD, standard deviation.

Supplementary Table 5. Summary of long COVID definitions used in all included SLR studies from both the initial and updated searches

| **Study** | **Time after SARS-CoV-2 infection** | **Diagnosis** | **Definition** |
| --- | --- | --- | --- |
| **Original SLR studies (up to March 2024)** | | | |
| AlBahrani 2023 | ≥3 months | Self-reported | NR, but study cites WHO consensus definition in Discussion: probable or confirmed SARS-CoV-2 infection with symptoms usually starting around 3 months from COVID-19 onset, persisting for at least 2 months, and not explained by an alternative diagnosis. |
| Antonelli 2023 | ≥4 weeks (≥12 week threshold also included) | Self-reported | Persisting pre-specified symptoms |
| Brown 2023 | 12 weeks | Self-reported | Ongoing symptoms and moderate-to-severe functional limitation. |
| Congdon 2023 | 4 months | Self-reported | Any of eleven listed common long COVID symptoms: dyspnoea, parosmia/dysgeusia, headache, dizziness, chest pain/tightness, palpitations, generalised fatigue, activity intolerance, nausea, vomiting or abdominal pain, brain fog, paraesthesia. |
| Di Fusco 2023 | 6 months (≥4 week is Sun 2024 secondary analysis) | Self-reported | ≥3 symptoms of 20 pre-specified symptoms based on the 2021 CDC list. |
| Diexer 2023 | ≥12 weeks | Self-reported | Presence of any of 24 symptoms: fever, swollen lymph nodes, smell and taste disorder, joint, muscle or limb pain, tiredness or fatigue (with normal adequate sleep duration), sleep disorder, cognitive impairment (concentration difficulties, memory loss, confusion), anxiety, depression, headache, congested nose, eye conjunctivitis, ear pain or ringing in the ears, shortness of breath, sore throat, cough, chest tightness/chest pain, heart complaints e.g., heart stuttering or cardiac arrhythmias, vertigo, abdominal pain, gastrointestinal complaints/diarrhea, nausea and vomiting, premenstrual syndrome/menstrual cramps. |
| Gallant 2023 | ≥1 month | Self-reported | Persistent symptoms captured using a 28-symptom questionnaire. |
| Hammel 2023 | ≥4 weeks | ICD codes | ICD 10 codes including: U.09.9, U07.1, Z86.16, and/or J12.82. |
| Herting 2023 | ≥4 weeks | Self-reported | Long-lasting symptoms |
| Li 2023 | ≥3 months | Caregiver-reported | One or more persistent physical symptoms for a minimum duration of 3 months with an impact on everyday functioning. |
| Lundberg-Morris 2023 | ≥28 days | ICD codes | U09.9 as the main or secondary diagnosis . |
| Mazzitelli 2023 | 1 month (3 month threshold also included) | Self-reported | Post-acute COVID-19 based on literature definitions. |
| Mikolajczyk 2023 | ≥4 months | Self-reported | Presence of post-COVID condition (from list of 21 symptoms). |
| Spiliopoulos 2023 | 4 months | Self-reported | The presence of ≥1 of symptoms listed (fatigue, concentration difficulties, headache, insomnia, smell issues, taste issues, dyspnoea, cough, myalgia, arthralgia, paraesthesia, chest pain, palpitations, digestive issues, hair loss). Only symptoms with a new onset after the test date were taken into consideration. |
| Richard 2023 | ≥1 month | Self-reported | New or increased difficulty exercising, new or increased difficulty doing daily activities (like walking or going up stairs). |
| Wander 2023 | ≥30 days | ICD codes | U09.9 for post–COVID-19 condition, unspecified |
| Woldegiorgis 2023 | 90 days | Self-reported | The presence of new or ongoing COVID-19 illness-related symptoms or health issues. |
| Huh 2024 | ≥30 days | Health care claims data | Occurrence of 26 conditions across eight domains, including cardiovascular and venous  thromboembolism, neurological, renal, respiratory, gastrointestinal, hepatic, and endocrine diseases. |
| Razzaghi 2024 | ≥28 days | ICD codes | Symptom-based or diagnosed long COVID (a single diagnosis code fell within the case definition for probable long COVID), or diagnosed long COVID only (two or more healthcare visits with diagnosis codes specific for long COVID). |
| Wu 2024 | ≥28 days | ICD codes | Conclusive or probable post-acute sequalae of COVID (a single healthcare visit indicating a diagnosis of PASC or MIS or the presence of a documented SARS-CoV-2 infection alongside a minimum of two long-COVID-compatible diagnoses). |
| **New SLR studies (March 2024 to July 2026)** | | | |
| Babicki 2024 | 3 months | Self-reported | Persistent clinical symptoms: coughs, shortness of breath, fatigue, smell and/or taste disorders, hair loss, impaired concentration and memory (brain fog), chest pain, headaches, and musculoskeletal pain. |
| Hori 2024 | ≥3 months | Self-reported | Any of the following persistent (lasting longer than two months) symptoms: fever, cough, fatigue, sore throat, chest pain, anorexia, brain fog, diminished ability to concentrate, anosmia, ageusia, dyspnoea, hair loss, muscle weakness, palpitations, sleep disorder, rhinorrhoea, headache, joint pain and swelling, muscle ache, feeling nauseous/vomiting, abdominal pain, skin rash, eye‐related symptoms, dizziness, erectile dysfunction (men only), and menstrual changes (women only). |
| Iba 2024 & Iba 2025 | ≥2 months | Self-reported | Any of the following symptoms that emerged during or after the first SARS-CoV-2 infection: fever, cough, fatigue, sore throat, chest pain, anorexia, brain fog, difficulty concentrating, anosmia, ageusia, shortness of breath, hair loss, muscle weakness, palpitations, sleep disorder, rhinorrhoea, headache, joint pain and swelling, muscle aches, nausea/vomiting, abdominal pain, skin rash, eye-related symptoms, dizziness, erectile dysfunction (male only), and menstrual change (female only). |
| Malden 2024 | ≥30 days | ICD codes | Any new-onset pre-specified outcome from the list of 13 post-COVID condition categories and 51 sub-conditions using ICD-10 codes for the following: endocrine and metabolic disorders, circulatory system disorders, blood and haematologic disorders, respiratory disorders, mental disorders, neurological or nervous system disorders, renal disorders, gastrointestinal disorders, symptoms (headache, body ache/myalgia, fever/malaise/fatigue, lymphadenopathy, weight loss, vertigo), skin and subcutaneous tissue disorders, sensory disorders, sleep disorders, and post-COVID-19 condition (unspecified). |
| Mohr 2024 | 6 weeks | Self-reported | Any symptom of the following of at least moderate severity (according to self-report): general symptoms (chills, difficulty with exercise, fatigue, fever), respiratory symptoms (cough, shortness of breath, severe respiratory illness, sore throat, cardiac symptoms (chest pain), neurologic symptoms (concentration difficulty, confusion, dizziness, headache, loss of taste/smell, memory difficulty, movement problems), gastrointestinal symptoms (abdominal pain, diarrhoea, nausea, vomiting), psychiatric symptoms (anxiety/panic, depression, trouble sleeping). |
| Omori 2024 | ≥30 days | Self-reported | Any of the following symptoms: cough, sputum, dyspnoea, chest discomfort, fever, musculoskeletal pain, headache, throat pain, loss of taste, loss of smell, cognitive impairment, insomnia, fatigue, muscle weakness, diarrhoea, and hair loss. |
| Xie 2024 | ≥30 days | ICD-10 codes, diagnosis, laboratory  values, and prescription medication codes | Any of the following symptoms: cardiovascular, coagulation and hematologic, fatigue, gastrointestinal, kidney, mental health, metabolic, musculoskeletal, neurological, and pulmonary. |
| Zheng 2024 | ≥2 months | Self-reported | Any of the following: fatigue, shortness of breath, chest pain or tightness, problems with memory and concentration (brain fog), difficulty to fall asleep, heart palpitations, dizziness, pins-and-needles, joint pain, feeling depressed or anxious, persistent cough, muscle pain, loss of smell or taste, and fever. |
| Butzin-Dozier 2025 | ≥1 month | ICD-10 codes | U09.9 (post-acute sequalae of COVID-19) |
| Carazo 2025 | ≥12 weeks | Self-reported | Any post-COVID-19 symptom(s) among a list of 21 (unreported) attributed to COVID-19. Moderate or severe long COVID was defined by the presence of ≥1 persistent moderate or severe symptom(s). |
| Caspersen 2025 | ≥3 months | Self-reported | Any of the following symptoms: cardiorespiratory (chest pain, cough, dyspnoea, heart palpitations, myocarditis, reduced lung function), neurocognitive (anxiety, brain fog, depression, dizziness, fatigue, headache, mood swings, poor memory, sleep problems), joint and muscle (joint pain, muscle pain), other (altered smell or taste, fever, hair loss, kidney disease, skin rash). |
| Liu 2025 | ≥6 months | ICD-10 codes | U09.9/B94.8, G93.31/R53.82 (post-acute sequalae of COVID-19) |
| Lutz 2025 | 3 months | Self-reported and EHRs | Presence of any self-reported symptom (new or persistent) that the participant thought to be related to their acute COVID-19 illness at the 3-month visit, any signs and symptoms of COVID-19 recorded in the EHR after the acute illness and up to the 3-month visit, and/or new report of one of the following medical conditions in the EHR after the acute illness and up to the 3-month visit: autoimmune condition, cardiovascular disease, cardiac dysrhythmia, heart failure, acute myocardial infarction, acute pulmonary embolism, asthma, renal failure (dialysis), chronic kidney disease, coagulation and haemorrhagic event, thromboembolic event, cerebrovascular event, gastrointestinal and oesophageal event, neurologic conditions, sleeping disorders, mental health conditions, substance-related disorders, muscle disorder, diabetes types 1 and 2. |
| Rai 2025 | ≥4 weeks (≥12 weeks threshold also included) | Diagnostic or referral clinical code | ≥1 long COVID diagnostic or referral clinical code in primary care , or ≥1 primary care clinical code for long COVID. |
| Sujarwoto 2025 | ≥12 weeks | Self-reported | Any of the following: fever, headache, muscle ache, weakness/tiredness, nausea/vomiting, abdominal pain, diarrhoea, loss of appetite, loss of taste, loss of smell, sore throat, cough, shortness of breath, chest pain, palpitations, vertigo/dizziness, worry/anxiety, low mood, trouble sleeping, memory loss, confusion, difficulty concentrating, runny nose/sneezing, wheezing. |
| Yadaw 2025 | ≥90 days | ICD-10 codes | ICD-10 U09.9, with additional potential long COVID patients identified using a machine learning (RECOVER ML) model2, ICD-10 code B94.8, or observation concept ID 2004207791. |
| Yousaf 2025 | ≥1 month | Self-reported | Any new or ongoing symptoms lasting at least 1 month after the positive SARS-CoV-2 test. |
| Lin 2026 | ≥8 weeks | Self-reported | Presence of any self-reported post-acute sequalae of COVID-19 symptoms (seven category-specific questions covering musculoskeletal, respiratory, gastrointestinal, neurological, cardiovascular, and other domains). |
| Towachiraporn 2026 | ≥4 weeks | Self-reported | Persistent symptoms starting during acute COVID-19 illness, new-onset symptoms following a period of symptom alleviation, evolution of symptoms with the addition of new symptoms over time or worsening of pre-existing symptoms. All of these conditions manifested at least four weeks after the infection with SARS-CoV-2. |

CDC, Centers for Disease Control and Prevention; EHR, electronic health record; ICD, International Classification of Diseases; SLR, systematic literature review.

Supplementary Table 6. Control of confounding variables reported by all included SLR studies from both the initial and updated searches

| **Publications** | **Control of confounding variables** |
| --- | --- |
| **Original SLR publications (up to March 2024)** | |
| AlBahrani 2023 | NR. |
| Antonelli 2023 | Age, BMI, sex, frailty, Index of Multiple Deprivation. |
| Brown 2023 | Age, sex, First Nations status, vaccine recency at the time of testing (<6 months (referent) vs 6 or more months since last COVID-19 vaccine dose), Index of Relative Socioeconomic Advantage and Disadvantage Score. |
| Congdon 2023 | Female sex, obesity (BMI ≥ 30), elderly age (≥ 65 years old), smoking history, diabetes, lung disease, and mood disorder. |
| Di Fusco 2023* | Time, vaccination status and interaction of time by vaccination status, as well as covariates of participant pre-COVID-19 symptom onset score, sociodemographic characteristics (age, sex, regions, social vulnerability, race/ethnicity, high risk occupations), previously tested positive for COVID-19, severity of acute illness (number of symptoms reported on index date), and immunocompromised status. |
| Diexer 2023 | Sex, age, country of birth (Germany vs other), living in a city, federal state, education, and perceived severity of the acute infection. |
| Gallant 2023 | Age, sex, BMI, smoking status, comorbidities, acute COVID hospitalisation. |
| Hammel 2023 | Age, sex, race, ethnicity, urban versus rural residence, the number of primary care visits in the past 24 months (0–5, 6–11, ≥12). |
| Herting 2023 | Female sex, age (years), Charlson comorbidity index, ≥3 immunosuppressants, difficulty concentrating, dyspnoea. |
| Li 2023 | Age, gender, ethnicity, type of residence (as a proxy for socioeconomic status), presence of comorbidities, and prior SARS-CoV-2 infection. |
| Lundberg-Morris 2023 | 1) Age, sex.  2) Age, sex, comorbidities (diabetes and cardiovascular, respiratory, and psychiatric disease), number of healthcare contacts during 2019, education level, employment status, and region of birth. |
| Mazzitelli 2023 | No adjustments. |
| Mikolajczyk 2023 | Months since last vaccination, months since last infection, number of symptoms at acute infection, sex, age, and study center. |
| Spiliopoulos 2023 | Age, sex, BMI, self-reported chronic diseases from the baseline questionnaire, Charlson comorbidity index, healthcare occupation, vaccination status (timing of vaccination with second dose before testing for SARS-CoV-2), and the week of infection during the period of Omicron predominance. |
| Richard 2023 | Time since first SARS-CoV-2-positive test (or enrolment for those who were never SARS-CoV-2 positive), their SARS-CoV-2 status (negative, unvaccinated positive, fully vaccinated positive), sex, age group (18–29, 30–39, 40+ years of age), BMI category, Charlson Comorbidity Index score >0, and military service affiliation, as well as a random effect for participant. |
| Wander 2023 | Age, sex, race, ethnicity, urban or rural residence, Charlson Comorbidity Index, VA’s administrative regions. |
| Woldegiorgis 2023 | Sex, age group, area of residence, pre-existing health issues. |
| Huh 2024 | Sex, age, insurance plans for low-income individuals, underlying diseases, a CCI score of 5 or higher, past influenza vaccination history, severity of COVID-19, and past medical usage as covariates. |
| Razzaghi 2024 | Sex, ethnicity, health system, baseline utilization in the 3 years prior to cohort entry, the number of body systems affected in the same interval, and the presence of a condition defined in the Pediatric Medical Complexity Algorithm as either progressive or malignant. |
| Wu 2024 | Age, sex, race/ethnicity obesity status, a chronic condition indicator as defined by the Pediatric Medical Complexity Algorithm, pre-existing chronic conditions, healthcare utilization metrics including the number of inpatient and outpatient visits, emergency department visits, unique medications prescribed, the count of negative COVID-19 tests administered prior to the cohort entry. |
| **New SLR publications (March 2024 to July 2026)** | |
| Babicki 2024 | Age, gender, BMI, diabetes, hypertension, asthma, thyroid diseases, hyperlipidaemia, previous heart attack(s), coronary heart disease, the presence of at least one chronic disease, and severity of COVID-19. |
| Hori 2024 | Age, sex, educational status, BMI. comorbidities, hospitalization, and COVID‐19 vaccination 14 days before infection. |
| Iba 2024** | 1) Age, sex, medical conditions, BMI, severity of acute COVID-19, and vaccination before infection.  2) Age, sex, medical conditions, BMI, severity of acute COVID-19, vaccination before infection, household income and education level. |
| Iba 2025** | Age (continuous variable), sex, BMI (continuous variable), pre-existing medical conditions (dummy variables for each condition), household income, educational level, and severity of acute COVID-19. |
| Malden 2024 | Vaccine Safety Datalink site, date of SARS-CoV-2 positive test (±7 days), age (exact year), sex, severity of infection (hospital admission with COVID-19 diagnosis within 7 days of SARS-CoV-2 positive test), prior SARS-CoV-2 infection, race and ethnicity, Charlson comorbidity score, Medicaid status, influenza vaccination, and healthcare utilization in the year prior. |
| Mohr 2024 | Age group, race and ethnicity, days to follow-up (as some participants completed their survey after 6 weeks), job classification, and the presence of at least 2 comorbidities. |
| Omori 2024 | Age, sex, BMI, smoking status, comorbidities, working status, and number of SARS-CoV-2 vaccinations received at the time of response. |
| Sun 2024* | Time point. |
| Xie 2024 | Age, race (White, Black, and Other), self-reported sex, ADI, smoking status (current, former, and never), use of long-term care, estimated  glomerular filtration rate, systolic and diastolic blood pressure, body mass index, cancer, cardiovascular disease, chronic lung disease, coronary artery disease, dementia, diabetes, hyperlipidaemia, HIV, immune dysfunction, liver diseases and peripheral artery diseases, number of outpatient visits and hospital admissions, number of blood panel tests, number of medications received, number of Medicare outpatient visits and hospital admissions, number of immunizations, receipt of seasonal influenza vaccine, and the predicted probability of receipt of COVID-19 tests during each era. |
| Zheng 2024 | Age, sex, severity of symptoms following SARS-CoV-2 infection. |
| Butzin-Dozier 2025 | All baseline covariates: demographics, BMI, comorbidities, immunocompromised status, corticosteroids, depression, lung disease, hypertension, obesity, diabetes, smoking, asthma, healthcare utilisation rate, date of inclusion, site and common data model format. |
| Carazo 2025 | Age group, sex, region of residence, occupation, workplace, material and social deprivation index, number of comorbidities. |
| Caspersen 2025 | Age. |
| Liu 2025 | Age, sex, ethnicity, BMI, date of acute COVID infection, antiviral treatment, hospitalization, number of COVID infections, asthma, COPD, smoking status, diabetes, hypertension, depression. |
| Lutz 2025 | Age group, sex, presence of underlying medical condition, medical presentation at time of acute illness, and vaccination status at time of acute illness. |
| Rai 2025 | Age; sex, ethnicity; country of GP practice; Index of Multiple Deprivation (2019) quintile; smoking status; body mass index; Quan-Charlson Comorbidity Index score; frailty status; influenza vaccination status; pneumococcal vaccination status; index quarter, previous COVID-19 diagnosis status. |
| Sujarwoto 2025 | Age, sex, education, income, occupation, comorbidities. |
| Yadaw 2025 | Demographics, comorbidities. |
| Yousaf 2025 | Sex (female and male or not reported), age at infection (5-11 years and 12-17 years), number of SARS-CoV-2 symptoms at time of acute infection (0-3 and ≥4), self-rated baseline health (excellent or very good and good, fair, or poor) and time between SARS-CoV-2 infection and post-COVID condition survey completion (60-129 days, 130-315 days, and 316 days). |
| Lin 2026 | Age, sex, race/ethnicity, BMI, current smoking, residence, antiviral therapy during acute phase, prior SARS-CoV-2 infection, severity of acute infection, general physical health, comorbidities, current or recent pregnancy. |
| Towachiraporn 2026 | Sex, age, occupation (healthcare vs non-healthcare). |

ADI, Area Deprivation Index; BMI, body mass index; CCI, Charlson Comorbidity Index; COPD, chronic obstructive pulmonary disease; HIV, human immunodeficiency virus; NR, not reported; SLR, systematic literature review; VA, Veterans Affairs.
* Di Fusco 2023^5^ and Sun 2024^27^ report the same study.
** Iba 2024^23^ and Iba 2025^33^ report the same study.

Supplementary Table 7. Vaccination association with long COVID

| **Publication/preprint** | **Vaccine(s)** | **Population** | **Time from infection** | **Dose comparison** | **Measure** | **Risk (95% CI)** | ***P* value** |
| --- | --- | --- | --- | --- | --- | --- | --- |
| **Original SLR publications (up to March 2024)** | | | | | | | |
| AlBahrani 2023 | NR | All | ≥3 months | Per 1 unit increase in dose number | OR | 0.53 (0.18–1.56) | 0.25 |
| Antonelli 2023 | NR | All | ≥4 weeks | 3 vs 2 | OR | 1.01 (0.85–1.19) | 0.943 |
|  |  |  | ≥12 weeks | 3 vs 2 | OR | 0.77 (0.39–1.52) | 0.448 |
|  |  | 18–59 years | ≥4 weeks | 3 vs 2 | OR | 0.91 (0.73–1.12) | 0.368 |
|  |  |  | ≥12 weeks | 3 vs 2 | OR | 0.73 (0.29–1.86) | 0.511 |
|  |  | ≥60 years | ≥4 weeks | 3 vs 2 | OR | 1.10 (0.83–1.45) | 0.001*** |
| Brown 2023 | NR | All | 12 weeks | ≥3 vs 0–2 | OR | 0.73 (0.56–0.96) | 0.02 |
| Congdon 2023 | NR | All | 4 months | 2–3 vs 0 | OR | *0.91 (0.46–1.79)* | 0.78 |
| Di Fusco 2023* | BNT162b2 | All | 6 months | 2 vs 0 | OR | 0.60 (0.27–1.34) | 0.296 |
|  |  |  |  | 3 vs 0 | OR | 0.36 (0.15–0.87) | 0.019 |
|  |  |  |  | 3 vs 2 | OR | 0.59 (0.21–1.65) | 0.459 |
| Diexer 2023 | NR | All | ≥12 weeks | 3 vs 0 | OR | *1.19 (0.92–1.56)* | NR |
|  |  |  |  | 3 vs 1–2 | OR | *0.78 (0.64–0.93)* | NR |
| Gallant 2023 | mRNA | All | ≥1 month | Per 1 unit increase in dose number | RR | 0.92 (0.79–1.98) | 0.489 |
| Hammel 2023 | Mixed | All | ≥4 weeks | 2 vs 0 | HR | 0.82 (0.79–0.86) | NR |
|  |  |  |  | 3 vs 0 | HR | 0.72 (0.68–0.76) | NR |
| Herting 2023 | Mixed | All | ≥4 weeks | ≥3 vs 0 | OR | 2.94 (0.84–10.28) | 0.085 |
| Li 2023 | mRNA | All | ≥3 months | ≥2 vs 0 | OR | 0.65 (0.34–1.25) | 0.20 |
| Lundberg-Morris 2023 | Mixed | All | ≥28 days | 1–3 vs 0 | HR (partially adjusted) | 0.59 (0.50–0.69) | NR |
|  |  |  |  |  | HR (fully adjusted) | 0.59 (0.50–0.69) | <0.001 |
| Mazzitelli 2023 | NR | All | 1 month | NR vs 0 | OR | 0.63 (0.44–0.89) | 0.009 |
|  |  |  | 3 months | NR vs 0 | OR | 0.76 (0.51–1.13) | 0.175 |
| Mikolajczyk 2023 | NR | All | ≥4 months | ≥4 vs 3 | OR | 0.52 (0.41–0.61) | NR |
| Wander 2023 | Mixed | All | ≥30 days | 2 vs 0 | HR | 0.80 (0.78–0.83) | NR |
|  |  |  |  | 3 vs 0 | HR | 0.66 (0.64–0.69) | NR |
| Woldegiorgis 2023 | NR | All | 90 days | ≥4 vs 0–2 | RR | *0.63 (0.52– 0.77)* | NR |
|  |  |  |  | ≥4 vs 3 | RR | *0.71 (0.63–0.77)* | NR |
| Razzaghi 2024 | mRNA | 5–11 years | ≥28 days | ≥1 vs 0 | OR | 1.34 (1.13–1.59) | NR |
|  |  | 12–17 years | ≥28 days | ≥1 vs 0 | OR | 0.99 (0.73–1.34) | NR |
| Wu 2024 | BNT162b2 | 5–11 years | ≥28 days | ≥1 vs 0 | RR | 1.24 (0.92–1.66) | NR |
|  |  | 12–20 years | ≥28 days | ≥1 vs 0 | RR | 0.91 (0.69–1.19) | NR |
| **New SLR publications (March 2024 to July 2026)** | | | | | | | |
| Babicki 2024 | NR | All | 3 months | ≥2 vs <2 | OR | 0.83 (0.57–1.19) | 0.302 |
| Hori 2024 | BNT162b2, mRNA-1273 | All | ≥3 months | ≥1 vs 0 | OR | 0.79 (0.59–1.07) | 0.13 |
| Iba 2024** | NR | All | ≥2 months | ≥1 vs 0 | OR (partially adjusted) | 0.74 (0.59–0.92) | 0.01 |
|  |  |  |  |  | OR (fully adjusted) | 0.75 (0.60–0.95) | 0.02 |
| Iba 2025** | NR | All | ≥2 months | 1–2 vs 0 | OR | 0.91 (0.68–1.22) | 0.53 |
|  |  |  |  | ≥3 vs 0 | OR | 0.69 (0.53–0.90) | 0.01 |
|  |  | Women | ≥2 months | 1–2 vs 0 | OR | 0.80 (0.57–1.13) | 0.21 |
|  |  |  |  | ≥3 vs 0 | OR | 0.70 (0.51–0.95) | 0.02 |
|  |  | Men | ≥2 months | 1–2 vs 0 | OR | 1.35 (0.78–2.32) | 0.28 |
|  |  |  |  | ≥3 vs 0 | OR | 0.83 (0.50–1.37) | 0.46 |
| Mohr 2024 | BNT162b2, mRNA-1273 | All | 6 weeks | 3 vs 2 | OR | 0.51 (0.40–0.65) | NR |
|  |  |  |  |  | OR (persistent symptoms only) | 0.48 (0.36–0.63) | NR |
|  |  | Booster ≤16 weeks of symptom onset | 6 weeks | 3 vs 2 | OR | 0.56 (0.43–0.73) | NR |
| Sun 2024* | BNT162b2 | 2022 study cohort | 4 weeks–6 months | 3 vs 0–2 | OR | *0.58 (0.39*–*0.85)* | NR |
| Xie 2024 | NR | All | 30 days–1 year | ≥1 vs 0 | IRR | 0.45 (0.43–0.47) | NR |
| Zheng 2024 | BNT162b2 | All | ≥2 months | 3 vs 2 | OR | 0.68 (0.52–0.90) | 0.006 |
|  |  |  |  |  | OR (≥3 symptoms) | 0.80 (0.55–1.18) | 0.26 |
| Butzin-Dozier 2025 | NR | All | 1 month–12 months | ≥2 vs 0 | RR | 0.843 (0.811–0.876) | NR |
| Carazo 2025 | BNT162b2, mRNA-1273 | Retrospective cohort | ≥12 weeks | 3–5 vs 2 | OR | 0.54 (0.44–0.67) | NR |
|  |  |  |  | 3 vs 2 | OR | 0.56 (0.45–0.69) | NR |
| Liu 2025 | BNT162b2, mRNA-1273 | All | 6–12 months | 1 vs 0 | OR | 0.77 (0.63–0.96) | NR |
|  |  |  |  | 2 vs 0 | OR | 0.73 (0.59–0.92) | NR |
|  |  |  |  | 3 vs 0 | OR | 0.64 (0.44–1.00) | NR |
|  |  |  |  | 4 vs 0 | OR | 0.29 (0.06–1.49) | NR |
| Lutz 2025 | NR | All | 3 months | ≥2 vs <2 | RR | 0.56 (0.34–0.90) | NR |
| Rai 2025 | NR | All | ≥4 weeks | ≥1 vs 0**** | IRR | 0.96 (0.82–1.13) | NR |
|  |  |  | ≥8 weeks | ≥1 vs 0**** | IRR | 0.98 (0.83–1.16) | NR |
|  |  | High risk | ≥4 weeks | ≥1 vs 0**** | IRR | 0.90 (0.71–1.14) | NR |
|  |  |  | ≥8 weeks | ≥1 vs 0**** | IRR | 0.93 (0.74–1.17) | NR |
|  |  | Highest risk | ≥4 weeks | ≥1 vs 0**** | IRR | 1.07 (0.82–1.40) | NR |
|  |  |  | ≥8 weeks | ≥1 vs 0**** | IRR | 1.06 (0.80–1.42) | NR |
| Sujarwoto 2025 | mRNA | All | ≥12 weeks | ≥1 vs 0 | OR | 0.549 (0.511–0.610) | <0.001 |
|  |  |  |  | 2 vs 1 | OR | 0.420 (0.402–0.511) | <0.001 |
|  | BNT162b2 | All | ≥12 weeks | ≥1 vs 0 | OR | 0.220 (0.057–0.771) | 0.010 |
|  | mRNA-1273 | All | ≥12 weeks | ≥1 vs 0 | OR | 0.341 (0.067–0.887) | 0.011 |
|  | Ad26.COV2.S | All | ≥12 weeks | ≥1 vs 0 | OR | 1.303 (1.073–1.630) | 0.012 |
|  | ChAdOx1 | All | ≥12 weeks | ≥1 vs 0 | OR | 1.381 (1.065–1.787) | 0.014 |
| Yadaw 2025 | NR | With pre-existing rare disease | ≥90 days | ≥2 vs <2 | OR | 0.71 (0.66–0.77) | <0.001 |
|  |  | Without pre-existing rare disease | ≥90 days | ≥2 vs <2 | OR | 0.64 (0.61–0.66) | <0.001 |
| Yousaf 2025 | BNT162b2 | All | ≥1 month | ≥2 vs 0 | OR (≥1 symptom LC definition) | 0.43 (0.19–0.98) | NR |
|  |  |  |  |  | OR (≥2 symptoms LC definition) | 0.27 (0.10–0.69) | NR |
| Lin 2026 | NR | All | ≥8 weeks | ≥1 vs 0 | HR | 0.83 (0.70–0.98) | NR |
| Towachiraporn 2026 | mRNA | All | ≥4 weeks | ≥1 vs 0 | OR | 0.41 (0.21–0.81) | 0.003 |

Spiliopoulos 2023, Richard 2023, Huh 2024, Malden 2024, Omori 2024, and Caspersen 2025 reported risks of individual symptoms only.
Data in italics have had their direction adjusted to match that of other studies (risk of higher number of doses compared with lower number of doses/no doses).
CI< confidence interval; HR, hazard ratio; IRR, incidence rate ratio; LC, long COVID; OR, odds ratio; RR, relative risk; SLR, systematic literature review.
* Di Fusco 2023^5^ and Sun 2024^27^ report the same study.
** Iba 2024^23^ and Iba 2025^33^ report the same study.
*** Likely error in publication.
**** Seasonal vs no seasonal vaccination

Supplementary Table 8. Dose comparisons and odds ratios included in meta-analyses

| **Publication** | **Vaccinated vs unvaccinated ORs (95% CI)** | **Booster dose vs primary course vaccination ORs (95% CI)** | **Booster dose vs unvaccinated ORs (95% CI)** |
| --- | --- | --- | --- |
| **Original SLR publications (up to March 2024)** | | | |
| Antonelli 2023 | - | **3 vs 2**: 0.77 (0.39–1.52) | - |
| Congdon 2023 | **2–3 vs 0**: 0.91 (0.46–1.79) | - | - |
| Di Fusco 2023 | **2 vs 0:** 0.60 (0.27–1.34) | **3 vs 1–2**: 0.59 (0.21–1.65) | **3 vs 0**: 0.36 (0.15–0.87) |
| Diexer 2023 | **3 vs 0:** 1.19 (0.92–1.56) | **≥3 vs 1–2**: 0.78 (0.64–0.93) | **≥3 vs 0**: 1.19 (0.92–1.56) |
| Hammel 2023 | **2 vs 0**: 0.82 (0.79–0.86)* | - | **3 vs 0**: 0.71 (0.67–0.76)^†^ |
| Lundberg-Morris 2023 | **≥2 vs 0**: 0.59 (0.50–0.69)^‡^ | - | - |
| Mazzitelli 2023 | **NR**: 0.76 (0.51–1.13) | - | - |
| Wander 2023 | **2 vs 0**: 0.80 (0.77–0.83)^§^ | - | **3 vs 0**: 0.65 (0.63–0.68)^\|\|^ |
| **New SLR publications (March 2024 to July 2026)** | | | |
| Babicki 2024 | **≥2 vs <2**: 0.83 (0.57–1.19) | - | - |
| Hori 2024 | **≥1 vs 0**: 0.79 (0.59–1.07) | - | - |
| Mohr 2024 | - | **3 vs 2**: 0.51 (0.40–0.65) | - |
| Zheng 2024 | - | **3 vs 2**: 0.68 (0.52–0.90) | - |
| Carazo 2025 | - | **3–5 vs 2**: 0.54 (0.44–0.67) | - |
| Iba 2025 | **1–2 vs 0**: 0.91 (0.68–1.22) | - | **≥3 vs 0**: 0.69 (0.53–0.90) |
| Liu 2025 | **2 vs 0**: 0.73 (0.59–0.92) | - | **3–4 vs 0**: 0.64 (0.44–1.00) |
| Sujarwoto 2025 | **≥1 vs 0**: 0.55 (0.51–0.61) | - | - |
| Towachiraporn 2026 | **≥1 vs 0**: 0.41 (0.21–0.81) | - | - |

* Converted from HR of 0.82 (0.79–0.86)
† Converted from HR of 0.72 (0.68–0.76)
‡ Converted from HR of 0.80 (0.78–0.83)
§ Converted from HR of 0.59 (0.50–0.69)
|| Converted from HR of 0.66 (0.64–0.69)
Rationale and methods for conversion of HRs described previously.^42^
CI, confidence interval; HR, hazard ratio; OR, odds ratio; SLR, systematic literature review.

Supplementary Table 9. Vaccinated vs unvaccinated sensitivity analyses

| **Analysis** | **Pooled OR (95% CI)** | ***P* value** | **Q** | **Degrees of freedom** | **Q *P* value** | ***I*^2^** | **tau^2^** |
| --- | --- | --- | --- | --- | --- | --- | --- |
| **Main analysis** | 0.75 (0.67–0.84) | <0.0001 | 94.48 | 12 | <0.0001 | 87.3% | 0.022 (0.008–0.137) |
| *Sensitivity analyses* | | | | | | | |
| Booster dose sensitivity analysis* | 0.69 (0.62–0.76) | <0.0001 | 55.73 | 12 | <0.0001 | 78.5% | 0.015 (0.006–0.132) |
| Excluding potential population overlap | 0.74 (0.64–0.87) | 0.0002 | 85.05 | 11 | <0.0001 | 87.1% | 0.048 (0.009–0.180) |
| Excluding high acute COVID-19 | 0.76 (0.63–0.91) | 0.0033 | 86.96 | 8 | <0.0001 | 90.8% | 0.056 (0.011–0.248) |
| Excluding unadjusted analyses | 0.75 (0.67–0.84) | <0.0001 | 94.47 | 11 | <0.0001 | 88.4% | 0.023 (0.009–0.156) |
| Including children/adolescent-only studies^†^ | 0.76 (0.68–0.84) | <0.0001 | 100.12 | 16 | <0.0001 | 84.0% | 0.022 (0.007–0.115) |
| *Leave-one-out analyses* | | | | | | | |
| Omitting Congdon 2023 | 0.75 (0.67–0.83) | - | - | - | - | - | - |
| Omitting Di Fusco 2023 | 0.75 (0.67–0.84) | - | - | - | - | - | - |
| Omitting Diexer 2023 | 0.72 (0.64–0.81) | - | - | - | - | - | - |
| Omitting Hammel 2023 | 0.74 (0.64–0.87) | - | - | - | - | - | - |
| Omitting Lundberg-Morris 2023 | 0.77 (0.69–0.86) | - | - | - | - | - | - |
| Omitting Mazzitelli 2023 | 0.75 (0.67–0.84) | - | - | - | - | - | - |
| Omitting Wander 2023 | 0.75 (0.63–0.88) | - | - | - | - | - | - |
| Omitting Babicki 2024 | 0.74 (0.66–0.84) | - | - | - | - | - | - |
| Omitting Hori 2024 | 0.75 (0.66–0.84) | - | - | - | - | - | - |
| Omitting Iba 2025 | 0.74 (0.66–0.83) | - | - | - | - | - | - |
| Omitting Liu 2025 | 0.75 (0.67–0.85) | - | - | - | - | - | - |
| Omitting Sujarwoto 2025 | 0.79 (0.73–0.85) | - | - | - | - | - | - |
| Omitting Towachiraporn 2026 | 0.76 (0.67–0.84) | - | - | - | - | - | - |

* In five studies (Di Fusco 2023,^5^ Hammel 2023,^8^ Wander 2023,^16^ Iba 2025,^33^ Liu 2025^34^) that reported both primary course vs unvaccinated and booster dose vs unvaccinated estimates, the estimate for the primary course was used in the main analysis, and replaced with the booster dose estimate in a sensitivity analysis.
† Inclusion of Li 2023,^10^ Razzaghi 2024,^19^ Wu 2024,^20^ and Yousaf 2025.^39^
CI, confidence interval; OR, odds ratio.

Supplementary Table 10. Booster dose vs primary course vaccination sensitivity analyses

| **Analysis** | **Pooled OR (95% CI)** | ***P* value** | **Q** | **Degrees of freedom** | **Q *P* value** | ***I*^2^** | **tau^2^** |
| --- | --- | --- | --- | --- | --- | --- | --- |
| **Main analysis** | 0.63 (0.53–0.75) | < 0.0001 | 10.63 | 5 | 0.0592 | 53.0% | 0.023 (0.000–0.207) |
| *Leave-one-out analyses* | | | | | | | |
| Omitting Antonelli 2023 | 0.62 (0.51–0.75) | - | - | - | - | - | - |
| Omitting Di Fusco 2023 | 0.63 (0.52–0.76) | - | - | - | - | - | - |
| Omitting Diexer 2023 | 0.57 (0.50–0.65) | - | - | - | - | - | - |
| Omitting Carazo 2025 | 0.66 (0.54–0.81) | - | - | - | - | - | - |
| Omitting Mohr 2024 | 0.67 (0.55–0.80) | - | - | - | - | - | - |
| Omitting Zheng 2024 | 0.62 (0.49–0.77) | - | - | - | - | - | - |

CI, confidence interval; OR, odds ratio.

Supplementary Table 11. Booster dose vs unvaccinated sensitivity analyses

| **Analysis** | **Pooled OR (95% CI)** | ***P* value** | **Q** | **Degrees of freedom** | **Q *P* value** | ***I*^2^** | **tau^2^** |
| --- | --- | --- | --- | --- | --- | --- | --- |
| **Main analysis** | 0.73 (0.64–0.82) | <0.0001 | 28.68 | 5 | <0.0001 | 82.6% | 0.013 (0.005–0.396) |
| *Sensitivity analyses* | | | | | | | |
| Excluding potential population overlap | 0.72 (0.55–0.95) | 0.0203 | 21.69 | 4 | 0.0002 | 81.6% | 0.067 (0.007–0.954) |
| *Leave-one-out analyses* | | | | | | | |
| Omitting Di Fusco 2023 | 0.73 (0.65–0.83) | - | - | - | - | - | - |
| Omitting Diexer 2023 | 0.68 (0.62–0.74) | - | - | - | - | - | - |
| Omitting Hammel 2023 | 0.72 (0.55–0.95) | - | - | - | - | - | - |
| Omitting Wander 2023 | 0.75 (0.59–0.96) | - | - | - | - | - | - |
| Omitting Iba 2025 | 0.73 (0.64–0.84) | - | - | - | - | - | - |
| Omitting Liu 2025 | 0.73 (0.64–0.84) | - | - | - | - | - | - |

CI, confidence interval; OR, odds ratio.

Supplementary Table 12. GRADE certainty of evidence assessment

| **Meta-analysis** | **N studies** | **Study design** | **Risk of bias** | **Inconsistency** | **Indirectness** | **Imprecision** | **Dissemination bias** | **Other considerations** | **Pooled risk (95% CI)** | ***P* value** | **Certainty of Evidence (GRADE)** |
| --- | --- | --- | --- | --- | --- | --- | --- | --- | --- | --- | --- |
| Vaccinated vs unvaccinated | 13 | Observational | None | Serious^a^ | Serious^d^ | None | None | None | OR 0.75  (0.67–0.84) | <0.0001 | ꚚOOO  Very low |
| Booster dose vs unvaccinated | 6 | Observational | None | Serious^b^ | Serious^d^ | None | None | None | OR 0.73  (0.64–0.82) | <0.0001 | ꚚOOO |
| Booster dose vs primary course | 6 | Observational | None | Serious^c^ | Serious^d^ | None | None | None | OR 0.63  (0.53–0.75) | <0.0001 | ꚚOOO |

^a^ Lower grading due to substantial heterogeneity, *I*^2^=87.3%, *P*(Q) <0.001.
^b^ Lower grading due to substantial heterogeneity, *I*^2^=82.6%, *P*(Q) <0.001.
^c^ Lower grading due to substantial heterogeneity, *I*^2^=53.0%, *P*(Q) = 0.0592
^d^ Lower grading due to inconsistency in long COVID definitions and follow-up periods, different vaccines doses across studies, and likely variable time since vaccination.
CI, confidence interval; GRADE, Grading of Recommendations Assessment, Development, and Evaluation; OR, odds ratio.

### Supplementary figures

Supplementary Figure 1. PRISMA flow diagram

**Identification of new studies via databases and registers**

**Screening**

Records screened

(n = 3,829)

Records excluded

(n = 3,628)

Reports sought for retrieval

(n = 201)

Reports not retrieved

(n = 1)

Reports assessed for eligibility

(n = 200)

Studies included in previous version of review (n = 20*)

Reports of studies included in previous version of review (n = 20*)

**Previous studies**

**Included**

**Identification**

Records identified from databases (n = 14,654)**

Records removed *before screening*:

Duplicate records removed (n = 10,825)**

Reports excluded (n = 179):

- Pre-Omicron/no stratification by variant (n = 83)
- Non-EMA-authorised vaccines (n = 33)
- No/wrong outcomes (n = 41)
- Duplicate (n = 9)
- No/wrong comparison (n = 8)
- Wrong design/publication type (n = 5)

New publications included

(n = 21)

- Publications of new studies (n=20; representing 19 studies)
- Publications of previously identified studies (n=1)

Total studies included in review

(n = 39)

Total publications included in review

(n = 41)

* Only studies reporting vaccination association with risk of long COVID
** Update searches run from 01 Jan 2022 and deduplicated against original searches

Supplementary Figure 2. Funnel plot for the vaccinated vs unvaccinated meta-analysis

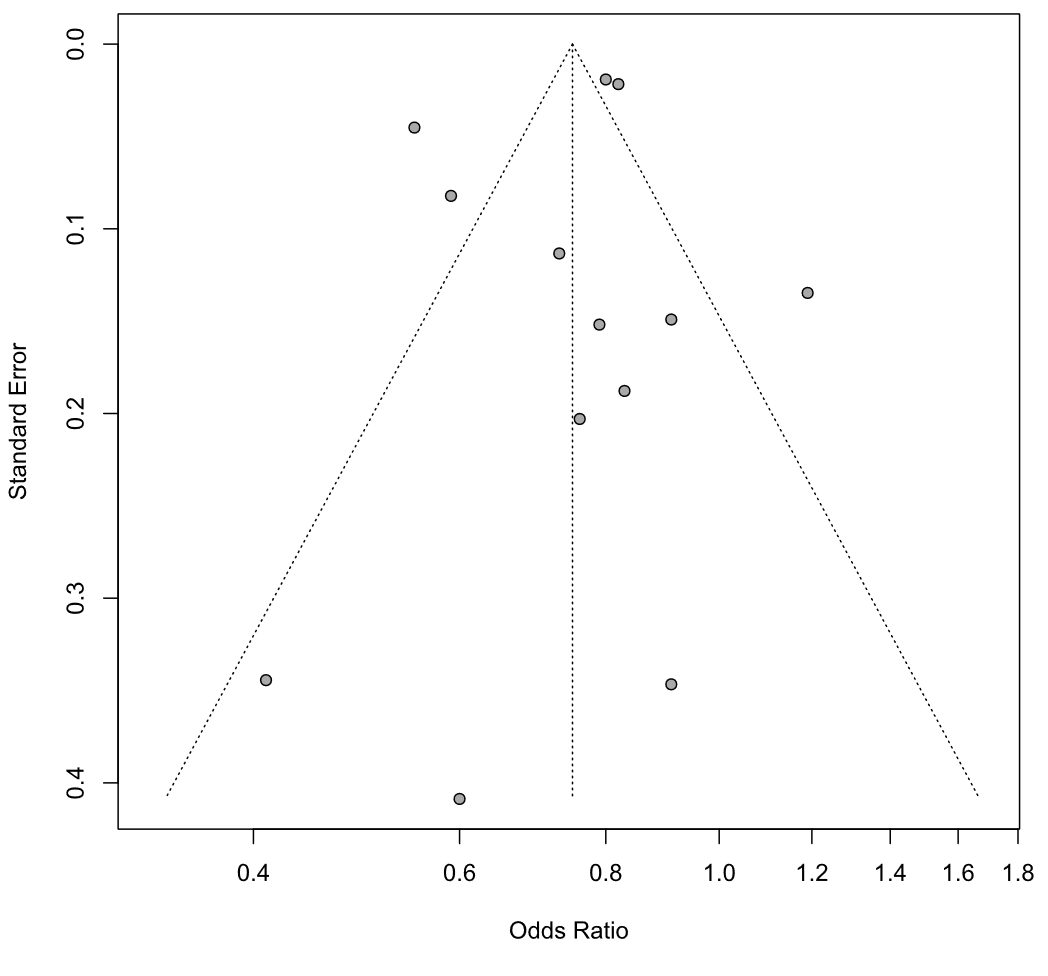

Supplementary Figure 3. Forest plot for the effect of COVID-19 booster doses on the risk of long COVID, compared with unvaccinated individuals

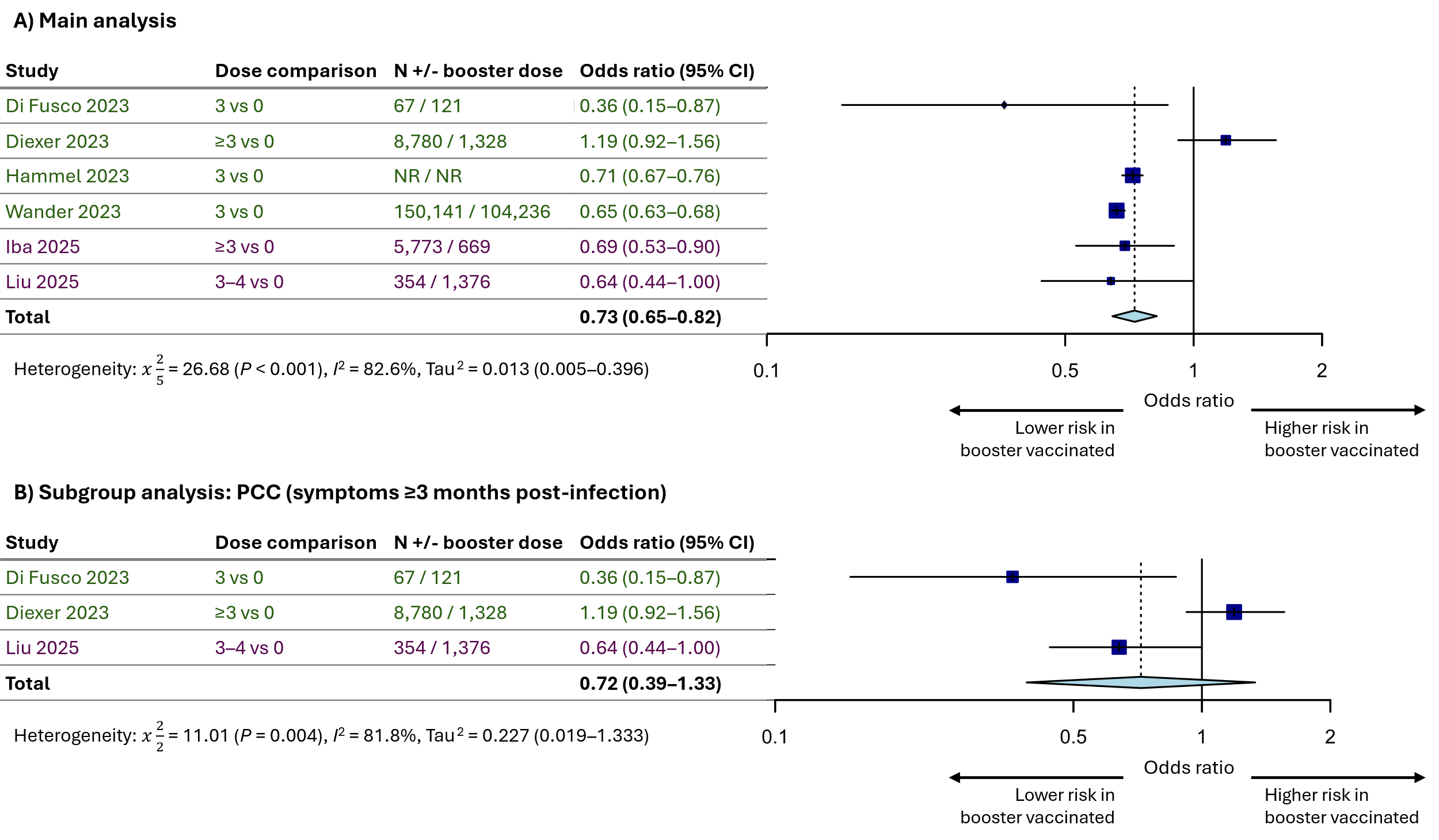

Original search studies = green; updated search studies = purple.
CI, confidence interval; PCC, post-COVID-19 condition.

Supplementary Figure 4. Forest plot for the effect of COVID-19 booster doses on the odds of long COVID, compared with individuals not receiving a booster dose

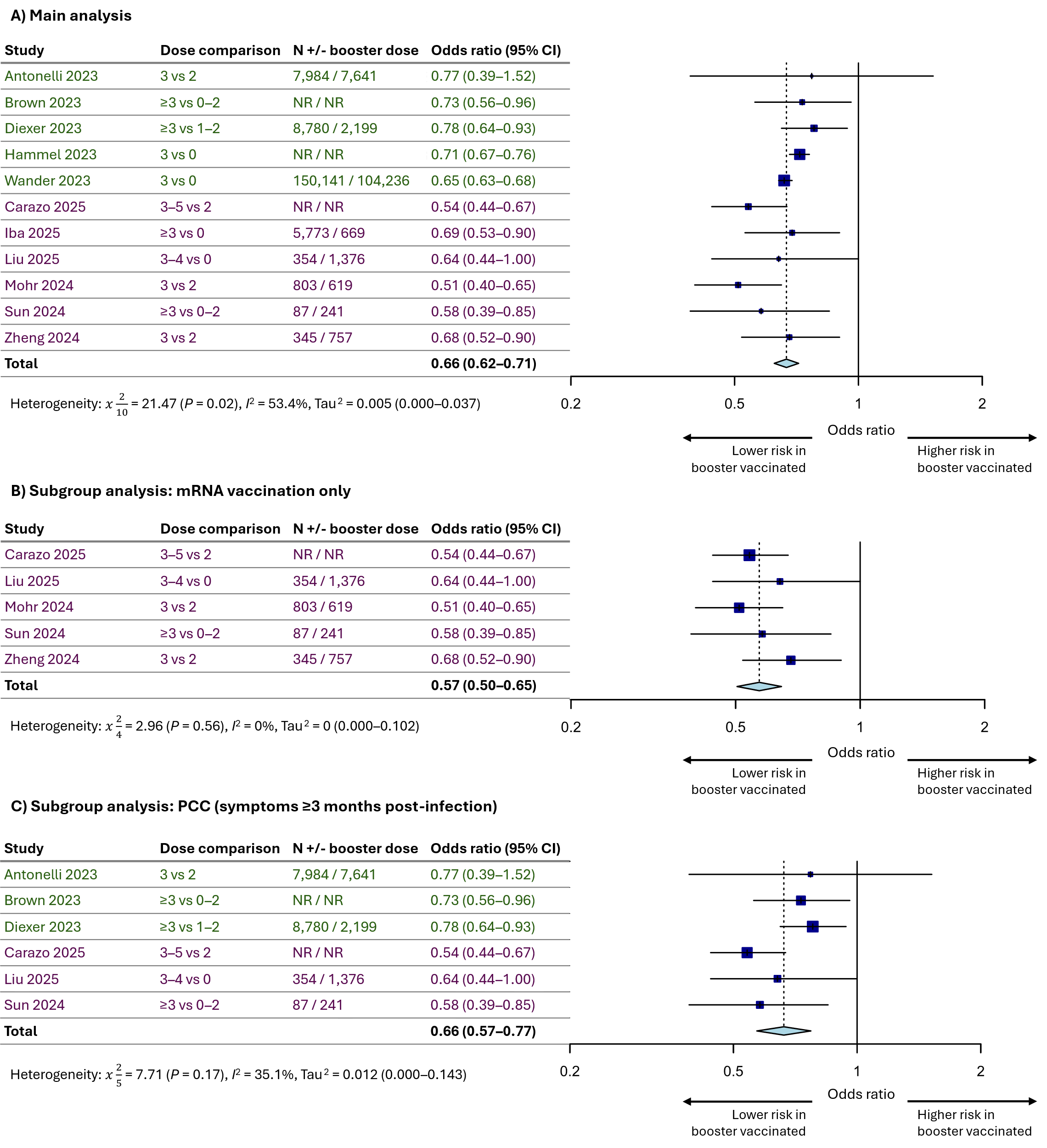

Original search studies = green; updated search studies = purple.
CI, confidence interval; PCC, post-COVID-19 condition.

### Meta-analysis feasibility assessment

The feasibility assessment aimed to determine the possibility of conducting pair-wise analyses of the association between pre-infection COVID-19 vaccination and the risk of developing long COVID after Omicron SARS-CoV-2 infection. The feasibility assessment combines all publications identified by the previously published systematic review,^42^ and all publications identified by the systematic review update.

The same methods used in the original systematic review feasibility assessment were used to assess publications, with the following changes: 1) pre-prints were excluded; 2) studies only including children and/or adolescents were excluded.

Publications were excluded from meta-analyses for the following reasons: individual long COVID symptoms reported only (six publications^14,15,18,24,26,32^); no comparable vaccine dose comparison (three publications^3,13,17^); same study as an included publication (two publications^23,27^); population was liver transplant patients only (one publication^9^); population was indigenous rural individuals in the USA only (one publication^35^); population was children/adolescents only (five publications^10,13,19,20,39^); publication was a pre-print (two publications^30,38^); there were fewer than 20 individuals in one comparison group (three publications^1,7,9^); reported a non-comparable risk measure (two publications^28,36^); reported HRs but incidence of long COVID was too high to be considered a rare (<10% event) and thus was not converted to ORs (one publication^40^).

### Sensitivity analyses

Sensitivity analyses performed (where possible) included:

- Vaccinated vs unvaccinated analysis only: replacement with booster dose comparison. In five studies reporting both vaccination dose comparisons of 2 vs 0 and 3 vs 0, the 2 vs 0 estimate was selected for the main analysis and replaced with the 3 vs 0 in the sensitivity analysis.
- Inclusion of adolescent and children-only studies
- Exclusion of potentially overlapping populations (removal of the study with the smaller population)
- Exclusion of studies with high acute COVID-19 illness hospitalisation (>5% of the population)
- Exclusion of studies reporting an unadjusted risk ratio.

A new sensitivity analysis was conducted post-hoc to capture the effect of booster vaccination compared with individuals who did not receive a booster dose. This analysis pooled any studies reporting a booster group compared with an unvaccinated, primary course-only, or a combined unvaccinated/primary course-only comparator (i.e., 0–2 doses). A comparison only of recent/seasonal vaccination versus no recent/seasonal vaccination was not possible due to only two studies reporting this comparison. Subgroup analyses of mRNA only vaccines and LC at ≥3 months post-infection were also performed. Studies and estimates included in this analysis are shown in Supplementary Table 13.

Supplementary Table 13. Dose comparisons and odds ratios included in the booster versus no booster sensitivity analysis

| **Publication** | **Booster dose vs no booster dose (unvaccinated and/or primary course only) ORs (95% CI)** |
| --- | --- |
| **Original SLR publications (up to March 2024)** | |
| Antonelli 2023 | **3 vs 2**: 0.77 (0.39–1.52) |
| Brown 2023 | **≥3 vs 0–2**: 0.73 (0.56–0.96) |
| Diexer 2023 | **≥3 vs 1–2**: 0.78 (0.64–0.93) |
| Hammel 2023 | **3 vs 0**: 0.71 (0.67–0.76)* |
| Wander 2023 | **3 vs 0**: 0.65 (0.63–0.68)^†^ |
| **New SLR publications (March 2024 to July 2026)** | |
| Mohr 2024 | **3 vs 2**: 0.51 (0.40–0.65) |
| Sun 2024 | **≥3 vs 0–2**: 0.58 (0.39–0.85) |
| Zheng 2024 | **3 vs 2**: 0.68 (0.52–0.90) |
| Carazo 2025 | **3–5 vs 2**: 0.54 (0.44–0.67) |
| Iba 2025 | **≥3 vs 0**: 0.69 (0.53–0.90) |
| Liu 2025 | **3–4 vs 0**: 0.64 (0.44–1.00) |

* Converted from HR of 0.72 (0.68, 0.76)
† Converted from HR of 0.66 (0.64, 0.69)
CI, confidence interval; OR, odds ratio; SLR, systematic literature review.
